# A Feed-Forward Loop Between Extrafollicular B Cell Differentiation and the Inflammatory Milieu Governs Remission and Relapse in Systemic Lupus Erythematosus

**DOI:** 10.64898/2026.07.03.26356448

**Authors:** Danae-Mona Nöthling, Kirill Anoshkin, Patrick G. Gavin, Tobias Rothe, Laura Bucci, Futoshi Iwata, Panagiotis Garantziotis, Nicola Ferrari, Sarah L.N. Clarke, Melanie Hagen, Andreas Wirsching, Jule Bachl, Carlo Tur, Sebastian Böltz, Sascha Kretschmann, Michael Aigner, Simon Völkl, Luis Munoz, Fabian Müller, Andreas Mackensen, Markus Eckstein, Maria Gabriella Raimondo, Aline Bozec, Georg Schett, Ricardo Grieshaber-Bouyer

## Abstract

Systemic lupus erythematosus (SLE) is driven by pathogenic B cells. Yet, why some patients receiving B cell depletion achieve durable remission, whilst others fail remains unclear. Here we use CD19-directed chimeric antigen receptor (CAR) T cell therapy as a mechanistic probe in 18 patients with refractory SLE^1,2^, with longitudinal follow-up extending up to 40 months. We show that durable, drug-free remission is defined not by the depth of B cell depletion alone, but by the elimination of the extrafollicular (EF) B cell differentiation trajectory – specifically, activated naïve B cell precursors and CD11c^+^ T-bet^+^ double-negative type 2 B cells. In long-term responders, B cell reconstitution recapitulated healthy ontogeny, while the EF pathway remained truncated, coinciding with collapse of the interferon-rich milieu and contraction of PD1^hi^ T peripheral helper cells. In contrast, in relapse, persistently elevated CXCL13, interferons and expanded PD1^hi^ T cells preceded the B cell return, and nascent B cells immediately followed the EF differentiation trajectory in the confirmed absence of germinal centers in the lymph node, shortly followed by clinical symptoms. These findings indicate that CAR-T cell therapy achieves remission by breaking a feed-forward loop between the systemic inflammatory environment and extrafollicular B cell differentiation.

## Introduction

Systemic lupus erythematosus (SLE) is a prototypical autoimmune disease characterized by loss of B cell tolerance, accumulation of autoreactive B cells and production of pathogenic autoantibodies directed against double stranded DNA and nuclear antigens. The disease predominantly affects young women and causes inflammation and tissue damage across organ systems^3^. Despite advances in management, remission induction fails in a substantial proportion of patients, and unmet needs regarding residual disease activity and treatment toxicity remain high^4,5^.

B cells in SLE are altered years before the disease clinically manifests^6,7^. The pathogenic B cells that sustain SLE are now understood to arise through two fundamentally different pathways. The classical model invokes defective germinal center (GC) selection, in which autoreactive clones escape negative selection and undergo somatic hypermutation and class-switching within organized lymphoid follicles^3,8–12^. However, accumulating evidence implicates an alternative, extrafollicular (EF) differentiation trajectory as a driver of active disease^13–15^. In this pathway, B cell receptor (BCR)-hyporesponsive activated naïve B cells bypass follicular checkpoints entirely and differentiate directly into antibody-secreting plasmablasts via a CD27^−^ IgD^−^ “double-negative type 2” (DN2) intermediate characterized by CD11c and T-bet expression^13,16–18^. This trajectory is further enhanced through exposure to autoantigens, TLR7 engagement and an interferon (IFN)-driven milieu seen in SLE^11,19–21^. Additionally, T peripheral helper (Tph) cells (PD1^hi^ CXCR5^−^ CD4^+^ T cells) are clonally expanded in inflamed tissues in the interferon-driven milieu of SLE and provide extrafollicular B cell help through secretion of IL-21 and CXCL13^22–24^.

While these EF-associated B cell populations are consistently expanded in active SLE and correlate with clinical flares^13,25–27^, their causal role in sustaining disease remains debated, as does their relationship to T cell help and the broader inflammatory environment. Hence, the presence of EF-associated B cell populations may maintain the pathologic immune landscape in SLE and thereby prevent curative treatment approaches. Conventional B cell depletion with anti-CD20 antibodies often fails to induce durable remission in SLE^28,29^, potentially because these agents incompletely deplete tissue-resident EF niches and EF intermediates rapidly re-emerge^30,31^.

This leaves open a fundamental question: Is autoimmunity perpetuated by a self-sustaining circuit linking the inflammatory environment to EF B cell differentiation or are intrinsic immune defects, such as genetic variants, dysregulation in TLR7 signaling, impaired BCR binding affinity^32,33^ and germline-encoded autoreactivity^15,34,35^ driving this process. In the latter model, B cell depletion would offer only temporary relief, as the regenerating compartment would inevitably recapitulate pathogenic differentiation (recently reviewed by Junt et al.^36^ and Zhu et al.^15^).

CD19-chimeric antigen receptor (CAR) T cell therapy offers a unique opportunity to resolve this dichotomy. By inducing deep, tissue-level depletion of B cells that exceeds the depth of monoclonal antibodies^37,38^, CAR-T cell therapy creates a natural experiment: the observation of B cell ontogeny de novo in adults whose inflamed milieu may, or may not, have been extinguished^1,2,39^. Here, we combine longitudinal high-dimensional immunophenotyping, multiplexed serum proteomics and single-cell RNA sequencing in cumulatively 18 patients with refractory SLE undergoing CD19-CAR-T cell therapy, with follow-up extending to more than 40 months. Collectively, we find that durable remission requires not merely deep B cell depletion in tissue, but the permanent abrogation of the pathogenic EF trajectory, and that relapse occurs in the context of a persistent inflammatory environment, which forces reconstituting B cells back into this maladaptive pathway.

## Results

### Extrafollicular B cell differentiation in refractory SLE

To define the immunological landscape of refractory SLE and establish a baseline for monitoring therapeutic response, we assembled a longitudinal cohort of 18 patients with multidrug-resistant SLE and 10 healthy controls (**Fig. 1a**, **Table 1**). All patients had active disease (median SLEDAI-2K: 15, range: 6–22), 83% had biopsy-confirmed lupus nephritis (LN) and the median number of prior therapies was 6. Follow-up after CAR-T cell therapy ranged from 3 to 40 months (median: 18 months, **Extended Fig. 1a**). Patients were systematically evaluated using an integrated platform encompassing high-dimensional spectral flow cytometry, multiplexed serum proteomics, single-cell RNA sequencing with paired B cell receptor profiling and detailed clinical assessments (**Fig. 1a**, **Extended Fig. 1b-g**).

**Fig. 1.**
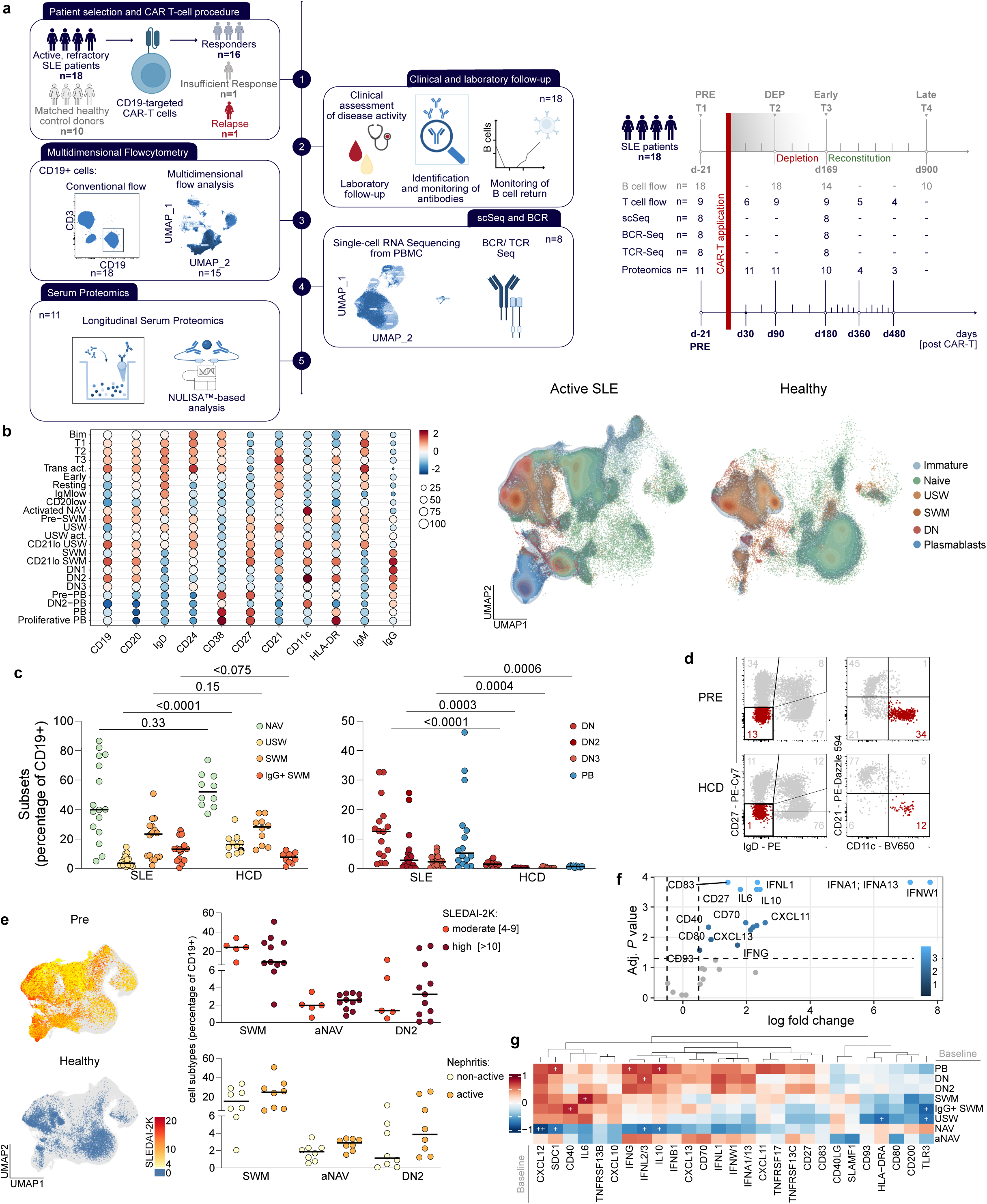
B cell dysregulation in active, treatment-refractory SLE. **a**, Study design of longitudinal immune-phenotyping of patients with active SLE patients undergoing CAR-T cell therapy. Results from n = 18 patients, of which n = 16 showed sustained response to treatment, n = 1 patient showed partial response and n = 1 patient relapsed. The patients included in each method shown out of n = 18 patients having received therapy. Median days (d) post treatment shown for landmark follow-up events. **b**, UMAP of B cells from patients (n = 16) with active disease and healthy control donors (n = 5) based on flow cytometry. Dot plots showing flow cytometry marker intensity. **c**, Distribution of B cell subsets (CD20^+^IgD^+^CD27^−^ NAV, CD20^+^IgD^+^CD27^+^ USW, CD20^+^IgD^−^CD27^+^ SWM, CD20^+^IgD^−^CD27^−^ DN, CD20^+^IgD^−^CD27^−^CD21^−^CD11c^+^ DN2, CD20^+^IgD^−^CD27^−^CD21^−^CD11c^−^ DN3, CD20^−^IgD^−^CD27^++^CD38^++^ PB); **Supplementary Fig. 1** shows gating strategy. Line indicates median. n = 10 - 16 per time point. Statistics: Kruskal-Wallis test with post hoc Dunn‘s multiple comparison test. **d**, Representative flow cytometry plots for DN2 B cells are shown from baseline and healthy. **e**, UMAP of B cells from flow cytometry data showing healthy donor (n = 5) cells and pre-treatment patients (n = 15) cells by SLEDAI-2K. Cell abundance is shown as percentage of CD19^+^ cells. Line indicates median. n = 10 - 16 per time point. Statistics: as in **c**. **f**, Baseline serum proteomic profile before therapy compared with healthy. Volcano plot showing log_2_ fold changes in baseline serum protein abundance between patients and healthy. Y axis shows p values; abundance assessed using Mann-Whitney U test followed by Benjamini-Hochberg correction; FDR < 0.05 considered significant. Proteins not significantly different after multiple-testing shown in grey. **g**, Heatmap showing Spearman correlation coefficients between baseline serum protein abundance and baseline frequencies of flow-cytometry-defined B cell subsets. Color indicates Spearman’s r. +, nominal p < 0.05; ++ Benjamini-Hochberg-adjusted p < 0.05.

**Table 1-.**
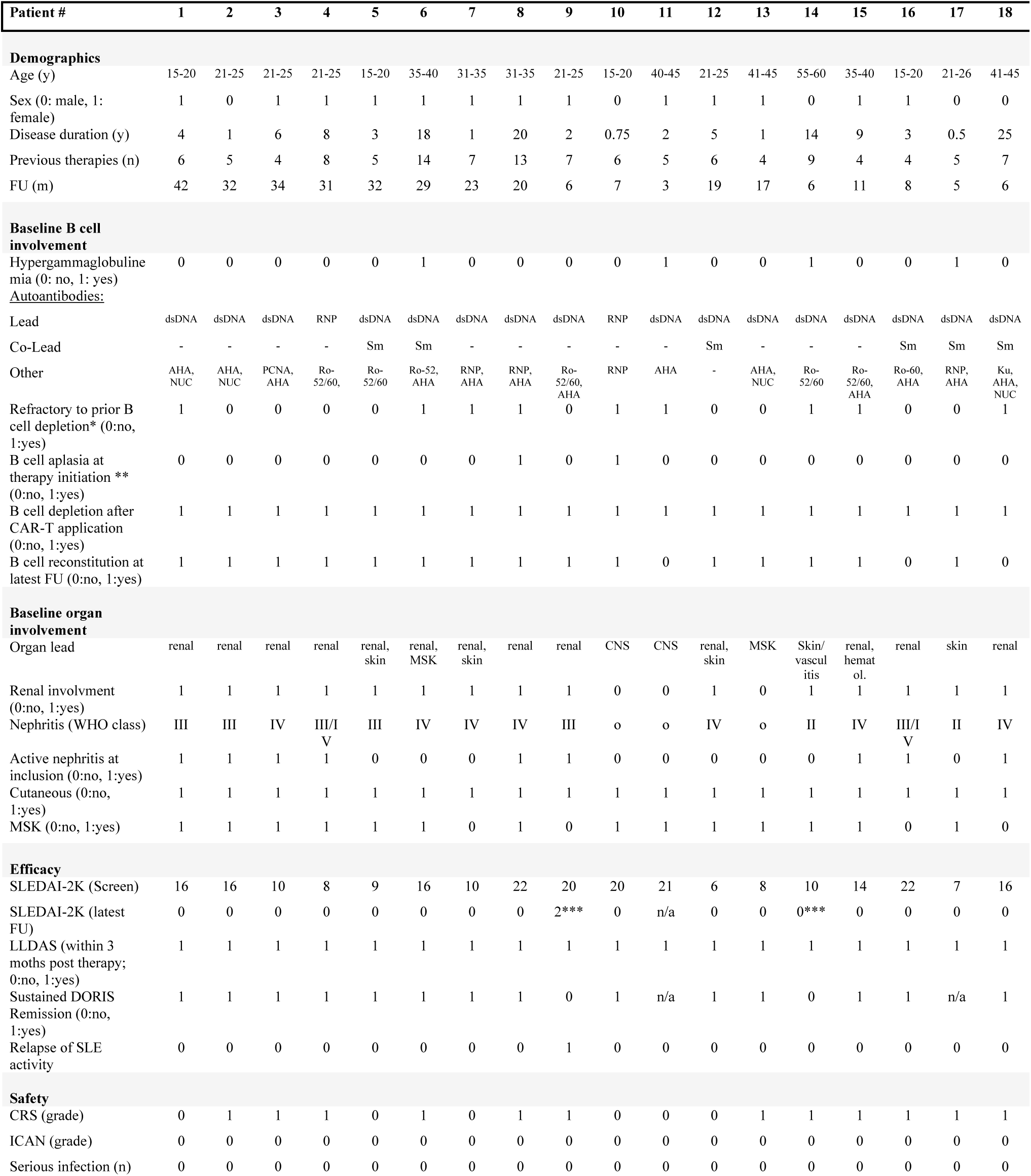
Clinical cohort characteristics. Baseline demographic, serological and clinical characteristics are shown for all 18 patients with SLE treated with CD19-CAR T-cell therapy. The table includes prior treatment exposure, baseline B-cell-associated disease features, organ involvement, treatment response and safety outcomes during follow-up. Sex is coded as 0, male and 1, female. Follow-up is reported in months from CAR T-cell infusion to the latest clinical assessment available. B cell depletion after CAR T-cell application refers to induction of peripheral CD19^+^ B cell aplasia after treatment. B cell reconstitution at latest follow-up refers to recovery of peripheral CD19^+^ B cells above the predefined reconstitution threshold (≥5 cells/µl). LLDAS (for patients with a minimum 3-month follow-up) and sustained DORIS remission at lasts follow-up were assessed according to established definitions. CRS and ICANS were graded according to standard consensus criteria. * Refractory to prior B cell depletion indicates previous exposure to B-cell-depleting therapy with insufficient clinical response, relapse after treatment or failure to achieve sustained disease control. ** B cell aplasia at therapy initiation indicates that peripheral CD19^+^ B cells were already absent or below the predefined detection/reconstitution threshold at the time of CAR T-cell therapy initiation, reflecting residual effects of prior B-cell-depleting therapy. *** The last SLEDAI-2K before follow-up cessation is shown; for relapsed patient #9, detailed SLEDAI-2K values are shown in **Fig. 5c**. **** For patient #14, the last value before follow-up cessation due to activity of hypocomplementemic vasculitis is shown.AHA, anti-histone antibodies; CNS, central nervous system; CRS, cytokine release syndrome; DORIS, Definition of Remission in SLE; FU, follow-up; ICANS, immune effector cell-associated neurotoxicity syndrome; LLDAS, lupus low disease activity state; MSK, musculoskeletal; n/a, not available or not assessable; NUC, anti-nucleosome antibodies; PCNA, proliferating cell nuclear antigen; SLEDAI-2K, Systemic Lupus Erythematosus Disease Activity Index 2000; WHO, World Health Organization.

Uniform Manifold Approximation and Projection (UMAP) analysis of high-dimensional flow cytometry data identified 23 discrete B cell clusters spanning transitional, naïve, double-negative (DN), memory and plasmablast (PB) compartments (**Fig. 1b,c**, **Extended Fig. 1d**; gating strategy for B cells is shown in **Supplementary Fig. 1**). At baseline, the most striking abnormalities compared to healthy controls were a 31-fold expansion of CD21^lo^ T-bet^+^ CD11c^+^ DN2 B cells (mean 5.71% vs. 0.18%), a 16-fold expansion of plasmablasts (11.13% vs. 0.69%) and a 13-fold expansion of DN3 cells (2.50% vs. 0.19%), accompanied by a reduction of unswitched memory and a trend to reduced naïve B cells (**Fig. 1c,d**). High disease activity (SLEDAI-2K ≥10) was associated with further enrichment of DN2 cells, and active LN showed a tendency toward additional expansion of activated naïve (aNAV) and DN2 populations relative to SLE patients without renal involvement (**Fig. 1e**).

Multiplexed serum proteomics identified a markedly elevated interferon signature encompassing type I (IFN-α, IFN-ω), type II (IFN-γ) and type III (IFN-λ1) interferons, alongside the interferon-inducible CXCR3-ligands CXCL10 and CXCL11, as well as CXCL13, which clearly separated patients with SLE from healthy controls (**Fig. 1f, Extended Fig. 1g**). Active LN was associated with a distinct proteomic signature characterized by further elevated IFN-λ1/2/3, BAFF-receptor (TNFRSF13C) and CD70, consistent with active B cell co-stimulation and extrafollicular class-switching (**Extended Fig. 1h**, **Supplementary Fig. 2**). Integration of cellular and proteomic data revealed coupling between the interferon-rich milieu and EF B cell expansion. IFN-λ2/3 levels in particular correlated with peripheral DN B cell counts during active disease, had no correlation with switched memory B cells and was negatively correlated with the number of naïve B cells. In addition, IFN-γ and sCD138 (SDC1) were correlated with the frequency of plasmablasts (**Fig. 1g**). CXCL13 as well as several IFN-induced chemokines including CXCL10 and CXCL11 were elevated in the serum of SLE patients, implicating engagement of T helper cells in active SLE (**Fig. 1f**, **Extended Fig. 1g**)^24^. Thus, active refractory SLE was not characterized by isolated expansion of DN2 cells, but by a co-occurring EF B cell and inflammatory state, linking DN/plasmablast expansion to interferons, CXCR3 ligands and CXCL13.

### Reset of B cell ontogeny via CAR-T cell therapy

Treatment with CD19-CAR-T cells induced rapid and complete depletion of circulating CD19^+^ B cells (median time to depletion: 0.5 days, IQR: 0-5.75; range of time to depletion: 0-9 days; **Supplementary Table 1**). B cell aplasia lasted a median of 137 days (IQR: 63-196). B cell reappearance (≥5 cells/µl) was observed in 15 of 18 patients (83%) during the observation time, and reconstitution to normal levels (≥60 cells/µl) was observed in 13 of 18 (72%) after a median of 213 days (IQR: 130.5-259) (**Fig. 2a**, **Extended Fig. 2**). Notably, elevated baseline serum TLR3 and IFN-λ showed a trend to delayed B cell reconstitution (**Extended Fig. 2g**) and higher baseline DN2 frequency independently predicted delayed reconstitution (p = 0.0475; **Extended Fig. 2f**). Active LN, characterized by the highest interferon burden, was additionally associated with prolonged time to B cell reappearance (**Extended Fig. 2d**). Together, these associations suggest that baseline EF and interferon-associated disease activity may influence the kinetics of B cell return, delaying B cell reconstitution in patients whose baseline B cell compartment shows stronger dependence on interferon-dependent and extrafollicular B cell differentiation.

**Fig. 2.**
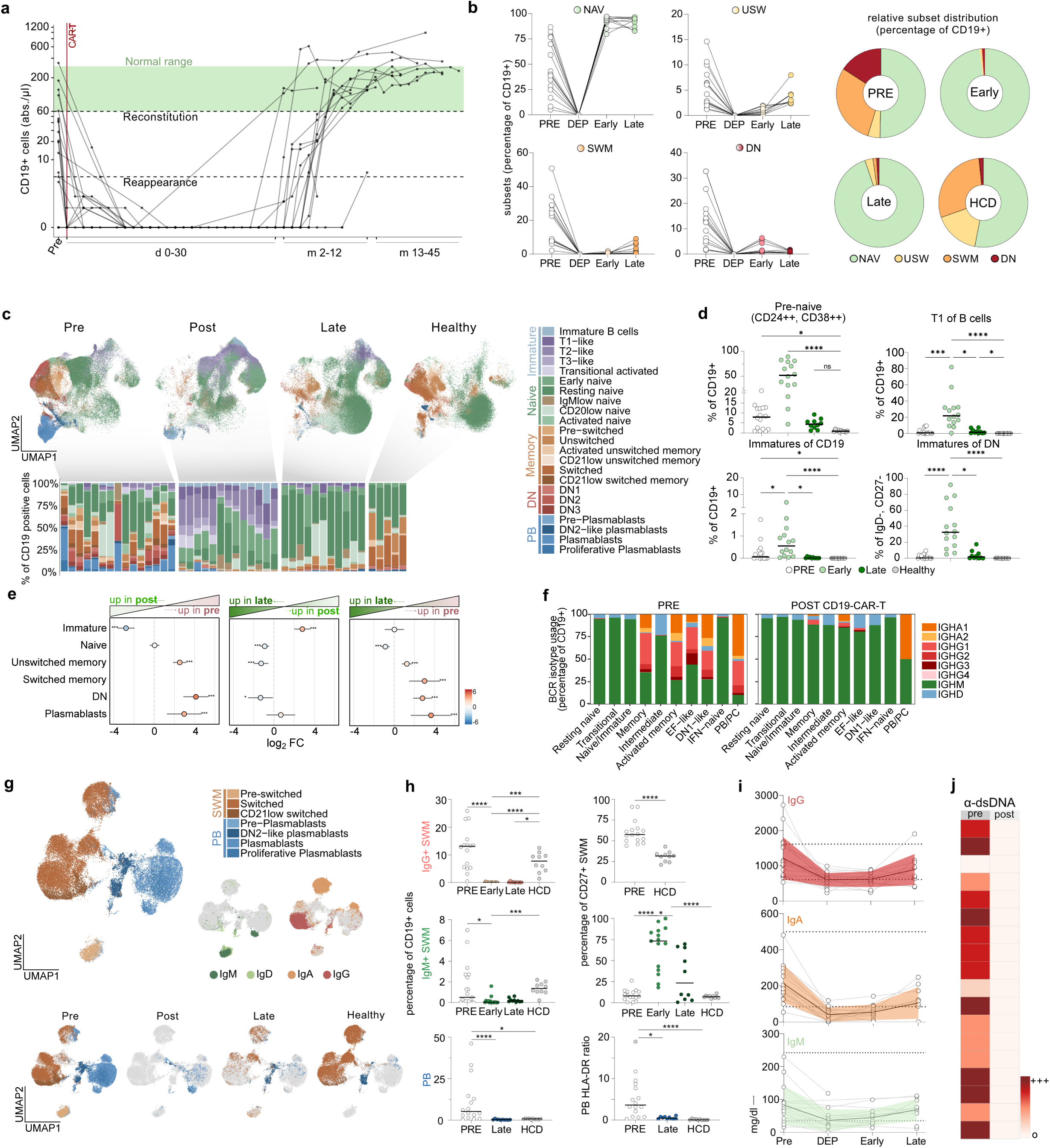
CD19-CAR-T cell therapy resets B cell ontogeny. **a**, Longitudinal absolute CD19^+^ B cells in blood from n = 18 patients following therapy. Time is shown in daily resolution for the first 30 days and in 30-day intervals thereafter. B cell reappearance (5 cells/µl) and reconstitution to reference levels (60 cells/µl) are indicated. Lines show median for intervals with multiple measurements. **b**, Frequencies of B cell subsets at baseline (Pre), depletion (Dep), early reappearance (Early), and late reconstitution (Late). Connected lines represent individual patients (n = 10 - 16 patients per timepoint). Part-of-a-whole plots show subset frequencies among CD19^+^ CD20^+^ B cells. **c**, UMAP and distribution of flow-cytometry-derived B cell phenotypes showing shifts in cluster composition during reconstitution. **d**, Frequencies of early B cell subsets by flow cytometry across timepoints. Violin plots show data distribution; lines indicate median and IQR (gating shown in **Supplementary Fig. 1**). n = 10 - 16 per timepoint. Statistics: Kruskal-Wallis test with post hoc Dunn‘s multiple comparison test. **e**, Changes in B cell subset abundance estimated by negative-binomial mixed-effects models with timepoint as fixed effect, patient as random intercept and offset for total cell counts. Points indicate model estimates ± 95% Wald CI. Statistics: pairwise Wald tests of estimated marginal means with Bonferroni adjustment. **f**, scRNA-seq-derived heavy chain isotype usage before and after therapy by B cell types. **g**, UMAP of switched memory B cells and plasmablasts showing cell type, surface immunoglobulin isotypes and timepoint. **h**, Quantification of memory B cell and plasmablast subsets over time and ratio of HLA-DR^hi^/HLA-DR^lo^ plasmablasts from flow cytometry. Statistics: as in **d**. **i**, Serum immunoglobulins over time. Individual trajectories (gray) are shown alongside the median (bold). Dotted lines and shading represent healthy reference range and IQR. **j**, Anti-dsDNA at baseline and latest follow-up (n = 18; semiquantitative values from negative (o) to highly positive (+++)).

B cell reconstitution recapitulated healthy ontogeny. The reconstituting compartment remained overwhelmingly naïve at all time points, even beyond 36 months post reconstitution (median: 94% of CD19^+^ cells; **Fig. 2b**). High-dimensional analysis further resolved B cell subsets longitudinally (**Fig. 2c**). Early reappearance was strongly dominated by CD24^hi^ CD38^hi^ CD10^+^ pre-naïve B cells, encompassing both CD21^−^ type 1 (T1) and CD21^+^ type 2 (T2) transitional subtypes, which accounted for up to 80% of circulating CD19^+^ cells (mean 50.35% vs. 0.97% in healthy controls; p < 0.0001; **Fig. 2c,d**). A distinct population of pre-transitional “immature B cells” (CD24^hi^ CD38^hi^ IgD^−^ IgM^hi^ CD10^+^ CD72^+^) was detectable at a mean frequency of 0.5% of CD19^+^ cells, far exceeding healthy donor levels. Of note, transitional and immature B cells were also modestly expanded in active SLE relative to healthy controls (p = 0.026 and p = 0.050, respectively), consistent with accelerated bone marrow output potentially driven by elevated IFN and BAFF levels^40,41^. Yet, their expansion at repopulation markedly exceeded this baseline elevation, in line with genuine ontogenic repopulation. Both populations declined progressively and returned to healthy donor levels at late follow-up, demarcating the completion of bone marrow repopulation (**Fig. 2d**). Those observed shifts in B cell family distribution remained apparent until late follow-up (**Fig. 2e**). Consistent with de novo B cell regeneration, BCR repertoire analysis showed a marked post-treatment shift toward IgM/IgD usage, reduced class-switched isotypes across B cell subsets (**Fig. 2f**).

Memory B cell- and plasmablast related serum proteins, including TACI (TNFRSF13B) and CD138 (SDC1), trended downwards after CAR-T cell therapy. This was observed even in patients without detectable B cells in circulation at therapy initiation (**Extended Fig. 3**), concordant with incomplete B cell depletion of prior CD20 mAb therapy and a deeper depletion in tissue achieved via CAR-T cell therapy^37^. Memory B cells were nearly absent at first B cell reappearance (median % of CD19^+^ cells: unswitched memory: 0.73; switched memory: 0.21) and expanded only gradually: IgM^+^ pre-switched memory B cells appeared first, while class-switched IgG^+^ memory B cells remained largely absent until late timepoints (**Fig. 2g,h**). Plasmablasts remained below 5% of CD19^+^ cells throughout follow-up, and the reconstituting switched memory B cells and plasmablasts displayed markedly reduced HLA-DR expression, with a significant decline in the HLA-DR^hi^/HLA-DR^lo^ ratio compared to baseline, consistent with durable elimination of the highly activated plasmablast phenotype characteristic of active SLE (**Fig. 2g,h**). Serum immunoglobulin levels decreased during B cell aplasia, resulting in mild hypogammaglobulinemia in 9/18 (50%) and reduced serum IgA in 15/18 (83.33%) of patients (**Fig. 2i**).

Higher disease activity as well as active nephritis at baseline were associated with development of hypogammaglobulinemia (**Extended Fig. 3**). Despite the only partial reduction of serum immunoglobulins, anti-dsDNA antibodies, present in 16 of 18 patients at baseline, became undetectable in all patients following therapy, confirming CD19^+^ plasmablasts and short-lived plasma cells as their cellular source **(Fig. 2j)**.

Together, these findings demonstrate that CD19 CAR-T cell therapy induces a durable reset of B cell ontogeny. Reconstitution proceeds through de novo differentiation along a naïve-dominant trajectory, with memory formation occurring gradually in a prototypical developmental sequence that is qualitatively distinct from the pathological baseline.

#### CAR-T truncates EF differentiation

Given the strong association between EF B cells and active SLE, it remains a central question whether reconstituting B cells will re-acquire a pathogenic phenotype, which could drive disease recurrence. To address this, we performed a comprehensive analysis of EF-associated phenotypes across all B cell subsets before and after CD19 CAR-T cell therapy. At baseline, sub clustering of naïve B cells revealed 10 distinct populations **(Fig. 3a)**. IgM^lo^ naïve IgD^+^ B cells [B_ND_] (first described in type 1 diabetes and linked to chronic antigen exposure^42^), were expanded compared to healthy donors (2.13% vs. 0.043%; p = 0.045 (median; % of naïve B cells; **Fig. 3b,c**). Within this pool, a CD21^lo^ CD11c^+^ subset (B_ND2_) displayed reduced CD24 and CD38 expression (**Fig. 3c**), as well as reduced expression of the immaturity marker CD10, compared to mature resting naïve B cells (p = 0.008; **Extended Fig. 4**), positioning B_ND2_ as an intermediate between naïve and DN B cells, consistent with a possible precursor of the EF differentiation axis. Transcriptional comparison of IgM^lo^ naïve B_ND_ cells provided further support for altered signaling within this compartment; IgD^only^ cells showed higher expression of genes linked to proximal BCR and PI3K/PKC signaling, including *VAV3*, *PIK3CA*, *PRKCB*, *PRKCE* and *RFTN1*, together with *ZEB2*, a master transcriptional regulator particularly linked to EF-like differentiation^43^. In contrast, IgM-expressing naïve B cells were enriched for BCR-complex and inhibitory checkpoint-associated genes, including *CD79A*, *CD79B*, *PTPN6/SHP1* and *FCGR2B* (**Supplementary Fig. 3**). Activated naïve B cells (aNAV; CD21^lo^ CD11c^+^) were similarly expanded (2.36% vs. 0.48%; p = 0.017 (median; % of CD19^+^ cells) and expressed reduced CD62L and elevated CD95, consistent with an interferon-driven activated phenotype (**Fig. 3d**, **Extended Fig. 5**). Concordantly, HLA-DR surface expression of reconstituting naïve B cells was significantly reduced compared to active disease (**Fig. 3e**, **Supplementary Fig. 4**), collectively indicating a shift from a chronically activated to a naïve state as a critical upstream checkpoint of immune reset. Together, these phenotypic and transcriptional features suggest that IgD^+^ IgM^lo^ naïve B cells and particularly B_ND2_ cells represent an activated intermediate of naïve B cells with remodeled signaling that may lie upstream of the autoreactive extrafollicular axis in active SLE.

**Fig. 3.**
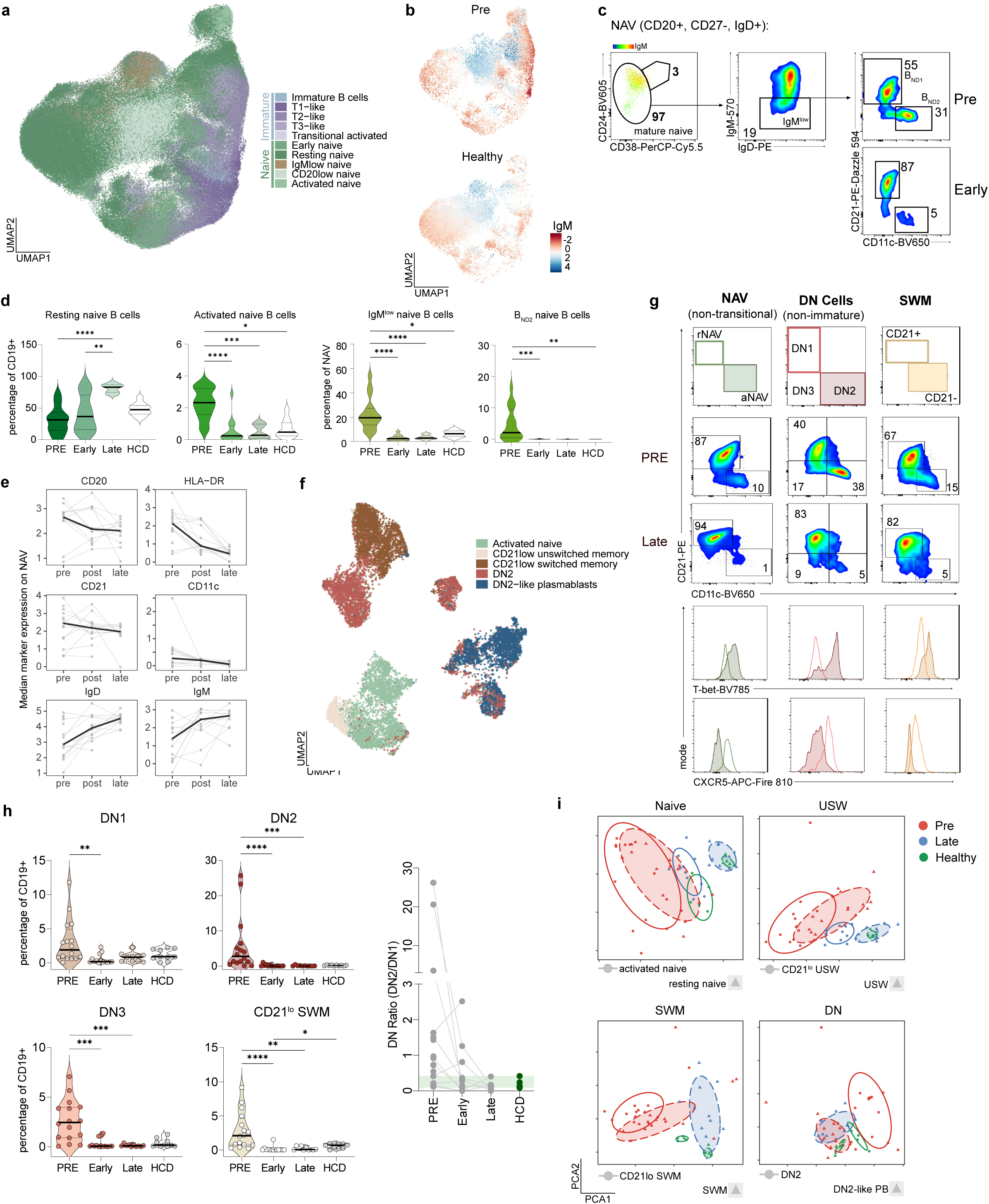
CD19-CAR-T cell therapy restores a naïve B cell compartment lacking extrafollicular features of active SLE. **a**, UMAP of naïve B cells from flow cytometry. **b**, UMAP showing surface IgM marker intensity in patients before therapy (n = 15) and in healthy controls (n = 5). **c**, Identification of CD24^++^CD38^++^IgM^low^ „IgM^low^“B cells. Within IgM^low^ B cells, B_ND1_ (CD21^+^ CD11c^−^) and B_ND2_ (CD21^−^ CD11c^+^) cells were identified. **d**, Frequencies of naïve B cell subsets are shown at predefined timepoints (baseline, depletion, early reconstitution, late reconstitution): resting naïve (IgD^+^CD27^−^CD24^−/low^), activated naïve (IgD^+^CD27^−^CD21^−^CD11c^+^), IgM^low^ naïve (IgD^+^CD27^−^IgM^lo/−^) and B_ND2_ (IgD^+^CD27^−^IgM^lo/−^CD21^−^CD11c^+^). Frequencies are relative to CD19^+^ cells or to naïve B cells, as indicated. n = 10 - 16 per timepoint. Statistics: Kruskal-Wallis test followed by Dunn’s multiple-comparison test. **e**, Longitudinal expression of selected surface markers within the naïve B cell compartment; black line: median. **f**, UMAP of atypical B cell subsets based on flow cytometry. **g**, EF-like subsets were defined by a CD21^−^CD11c^+^ surface phenotype within non-transitional naïve, double-negative and memory B cells. Gating from one patient is shown. EF-like phenotype was further confirmed by T-bet expression and loss of CXCR5, illustrated in the histograms. **h,** Frequencies of the indicated subpopulations were assessed by flow cytometry at timepoints as in **c**. Within the double-negative B cell compartment, subsets were defined as DN1 (CD20^+^IgD^−^CD27^−^CD21^+^CD11c^−^), DN2 (CD20^+^IgD^−^CD27^−^CD21^−^CD11c^+^) and DN3 (CD20^+^IgD^−^CD27^−^CD21^−^CD11c^−^). CD21^lo^ switched-memory (SWM) B cells were gated as CD20^+^IgD^−^CD27^+^CD21^−^CD11c^+^. n = 10-16 per timepoint. Statistics: as in **d**. The quotient ratio of DN2 per DN1 cells is shown per timepoint and patient. The range of healthy donor minimum to maximum is shown in green. **i**, Principal component analysis of median marker expression across B cell subclusters and time points. Points represent one sample within the indicated subcluster and time point. Colors denote disease or treatment state, symbols indicate cell subclusters and ellipses indicate the 68% confidence region for each subcluster-time group.

To map EF-associated phenotypes across all B cells, we applied a unified gating strategy based on CD11c expression and CD21 downregulation, revealing that EF-skewed phenotypes were distributed across naïve, memory and DN compartments (**Fig. 3f,g**). Within the DN compartment, three subpopulations were resolved: DN1 (CD21^+^ CD11c^−^), DN2 (CD21^−^ CD11c^+^ T-bet^hi^ CXCR5^lo^) and DN3 (CD21^−^ CD11c^−^).

To enhance the resolution of rare EF-like and other rare B and T cell populations in the post-depletion window, scRNA-seq analyses was performed. We integrated a publicly available cohort of patients treated with dual CD19/BCMA CAR-T cell therapy (PRJNA1303131) to further augment the representation of low-frequency cell states, as shown in analyses presenting results before (PRE) and after (POST) CAR-T cell therapy. Where cohort composition was relevant to interpretation, analyses were stratified and indicated separately; the relapsing patient was analyzed as an independent contrast trajectory throughout.

DN2 carried the canonical EF transcriptional signature as validated by scRNA-seq (**Extended Fig. 5a,c**), which also identified a subcluster of EF-like B cells containing both IgD/IgM-expressing naïve-leaning cells and class-switched DN2-like cells, consistent with a mixed cluster of naïve and memory EF-like in single-cell sequencing subsets analysis of BCR isotypes (**Extended Fig. 6a**).

Following CAR-T cell therapy, all EF-associated subsets were durably suppressed. B_ND2_ cells were completely abolished and remained undetectable at all subsequent timepoints. aNAV cells declined significantly, and those reappearing during reconstitution expressed normalized surface IgM levels. Long-term follow-up showed no enrichment of IgM^lo^ naïve B cells even beyond 36 months, consistent with the notion that these phenotypes reflect chronic antigen stimulation rather than a constitutive feature of B cell homeostasis (**Fig. 3h**). DN2 and DN3 frequencies remained at or below healthy donor levels throughout follow-up, without re-expansion in any patient maintaining durable remission (**Fig. 3h**). As memory slowly reconstituted, the DN compartment re-expanded preferentially through DN1, reflected in a normalized DN1/DN2 ratio (**Fig. 3h**). Only two patients had a small number of detectable DN2 cells at a late reconstitution time point. As B cells had previously been fully depleted, new DN2 were predominantly IgM^+^ IgG^−^ (>90%), in contrast to their IgG-dominant baseline phenotype. This observation is in line with nascent generation rather than re-emergence of existing clones (**Extended Fig. 4e**). Concordantly, BCR analysis showed reduced somatic hypermutation frequencies after therapy across B cell states and isotypes, including EF-like B cells, further supporting de novo generation during B cell reconstitution instead of re-expansion of residual antigen-experienced clones (**Extended Fig. 6**).

Global flow-cytometric surface marker analysis confirmed that late post-therapy B cell phenotypes shifted towards and overlapped with healthy donor profiles across all subsets, indicating broad phenotypic normalization (**Fig. 3i**, **Supplementary Fig. 5**).

Pseudotime analysis further indicated altered positioning of reconstituting B cells along the inferred B cell trajectory. After CAR-T cell therapy, EF-like B cells shifted towards lower pseudotime values and were positioned closer to naïve B cell states than at baseline (**Supplementary Fig. 6**). Timepoint-specific trajectory modelling identified 301 genes preferentially associated with the baseline (PRE) pseudotime trajectory, including *IFITM3* and *PYHIN1*, together with genes involved in cytoskeletal organization and translational activity (**Supplementary Fig. 7**). These findings suggest that B cell differentiation was accompanied by an inflammatory, IFN-conditioned activation program distinct from the post-treatment state.

In healthy controls, EF-like B cell subsets showed relatively high CD72 surface expression, indicating that acquisition of an activated or EF-like phenotype is normally accompanied by inhibitory checkpoint reinforcement (**Extended Fig. 4g**). In active SLE, however, CD72 expression was selectively reduced within EF-associated subsets, consistent with impaired BCR/TLR7 signal restraint in the pathogenic EF compartment. Such reduced CD72 was recently associated with higher disease activity and resistance to rituximab treatment^44^. After CAR-T cell therapy, surface CD72 expression increased, particularly in aNAV B cells. Consistent with this phenotypic normalization, scRNA-seq showed preservation of inhibitory checkpoint-associated transcripts, including *CD72*, *FCGR2B* and *INPP5D*, and an increased composite BCR inhibitory score. Concordantly, a decrease in a BCR activation score was seen particularly in EF-like B cells after therapy (**Extended Fig. 4f**). Thus, reconstituting B cells did not simply lack EF-associated features but showed reconstitution of a B cell compartment lacking the CD72-low, poorly restrained activation state characteristic of active SLE.

These data indicate that durable remission is characterized by the sustained truncation of the EF differentiation trajectory: naïve precursors are eliminated, downstream pathogenic intermediates (DN2, DN3, activated PB) fail to re-emerge, and reconstitution proceeds through healthy developmental pathways. This suggests that not only the B cell compartment itself, but also effects on the host environment, contribute to reconstitution without utilization of the EF pathway.

#### IFN and T cell contraction reset SLE

To understand why reconstituting B cells did not re-engage the EF pathway, we interrogated the systemic immune environment longitudinally. Time-resolved serum proteomics revealed that the interferon-dominant signature declined progressively after therapy, with most dysregulated proteins normalizing to the healthy range (± 2 SD) over 12 months (**Fig. 4a**). Notably, interferons and interferon-induced chemokines did not normalize abruptly but trended progressively towards healthy levels, reflecting gradual rebalancing rather than an immediate consequence of B cell removal.

**Fig. 4.**
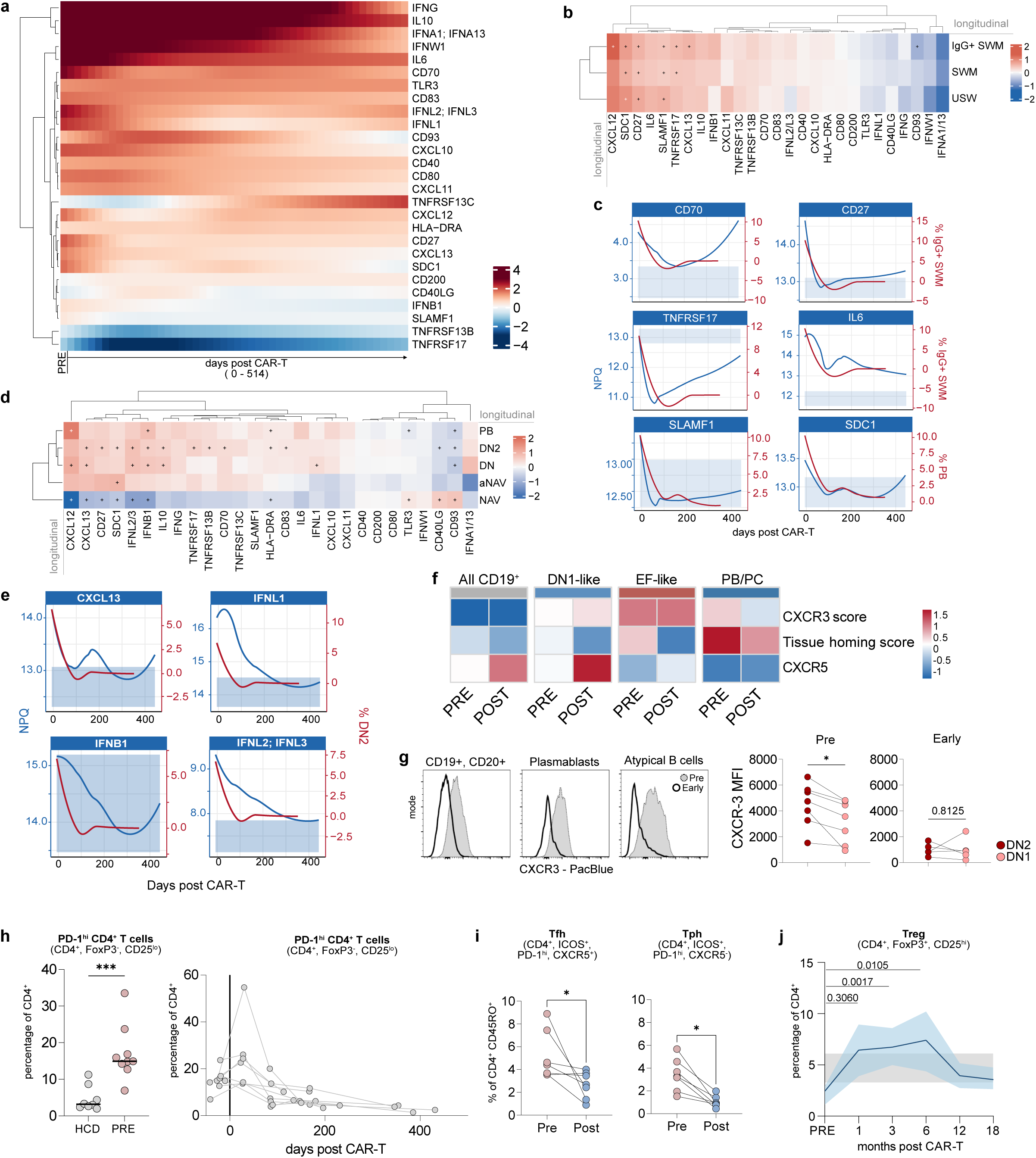
Collapse of the inflammatory environment after deep B cell depletion. **a**, Longitudinal serum proteomics after therapy. Heatmap shows selected interferon-associated and immune-regulatory serum proteins from baseline until 514d post treatment. Values indicate standard deviations from the healthy-control median. Rows were hierarchically clustered (n = 10). **b**, Time-adjusted longitudinal associations between serum proteins and memory B cell frequencies. Protein abundance was modeled as a function of non-linear time after therapy and B cell subset frequency, with patients as a random effect. Colors indicate the β-coefficient for the B cell term; Benjamini-Hochberg-adjusted p ≤ 0.05 is marked (*). **c**, Overlay of selected serum proteins and corresponding memory B cell subset frequencies after CAR-T cell therapy. Blue lines show serum protein abundance as normalized protein expression values (NPX). Red lines, B cell subset frequencies. Shaded blue, healthy-control mean ± s.e.m.. **d**, Longitudinal association of serum proteins with non-memory B cell subset abundance, as in **b**. **e**, Overlay of selected inflammatory and interferon-associated serum proteins with selected B cell frequencies from flow cytometry. Blue: Normalized protein levels (NPX). Red: DN2 frequencies from flow cytometry. **f,** Single-cell RNA-seq-derived mean scores for CXCR3-associated and tissue-homing programs across cell states before and after therapy. Score genes shown in **Supplementary Table 4**. **g**, Surface CXCR3 MFI was compared on indicated B cell subsets (n = 7). Left: Histograms from representative samples. Right: Paired samples of CXCR3 MFI on DN1 and DN2 pre and post therapy. Statistics: Wilcoxon signed-rank test. **h**, Left: Quantification of PD1^high^CD4^+^ cells within CD4^+^CD25l^ow^FoxP3^low^ cells in SLE and healthy. n = 9 - 10 per timepoint. Statistics: Mann-Whitney U test. Right: Longitudinal quantification after therapy (n = 9 individual patients). **i**, Paired quantification of circulating Tfh cells and Tph cells before and after therapy. Tfh: CD4⁺ICOS⁺PD-1⁺CXCR5⁺; Tph: CD4⁺ICOS⁺PD-1⁺CXCR5⁻ (n = 7; gating shown in **Supplementary Fig. 9**). Statistics: Wilcoxon signed-rank test. **j**, Frequency of Tegs amongst CD4^+^ cells from flow cytometry. Bold, median. Shaded, IQR (n = 9 per timepoint). Statistics: Kruskal-Wallis test, Dunn’s multiple-comparison test.

Longitudinal correlation analysis revealed biologically informative coupling patterns of B cell subsets with soluble markers. Using repeated-measures correlation to account for within-subject dependence, changes in the mean CD27, TNFRSF17 (BCMA), SLAMF1, and SDC1 (CD138) were strongly associated with concomitant changes in expanded memory B cell subsets (**Fig. 4b**). Time-resolved proteomics demonstrated a sharp and durable decline of mean of CD70, CD27, TNFRSF17 (BCMA), IL-6, SLAMF1 and SDC1 (CD138), paralleling the decrease in mean abundance of memory B cell subsets and indicating sustained attenuation of memory and plasmablast survival signals (**Fig. 4c**). Aligned with B cell reappearance, we observed CD27 and SDC1 tracking switched memory B cell frequencies, declining during depletion and remaining low at reappearance, while subsequent B cell return was associated with sharply rising TNFRSF13C (BAFF-R) and only gradual increase in TNFRSF13B (TACI). In contrast, CXCL10, CXCL11 and IL-10 continued to decline throughout follow-up, indicating continued immune remodeling even after the onset of B cell recovery **(Supplementary Fig. 2b)**.

With EF-like B cells having been deemed IFN-λ hyperresponsive^45^, type III interferons (IFN-λ2/3) correlated longitudinally with the frequency of DN2 cells (**Fig. 4d,e**). *IFNLR1* was expressed in the majority of EF-like B cells, whilst the plasmablast compartment showed decreased expression post therapy (**Supplementary Fig. 8a**). This implicates IFN-λ signaling as a potential driver of EF differentiation into plasmablasts during active disease and could support serum IFN-λ as potentially specific marker for EF-activation.

Memory B cell trajectories correlated predominantly with SLAMF1 and soluble BCMA, dissociating the EF-specific inflammatory program from the canonical memory B cell survival axis (**Fig. 4b,d**). Ligands for CXCR3 (CXCL10/11) additionally normalized over time (**Fig. 4a**); CXCL13 declined sharply in responders, indicating rapid collapse of the follicular chemokine program, and B cell-associated proteins (CD27, TACI, BAFF-R, SLAMF1) showed parallel durable suppression (**Fig. 4a,f**). After controlling for the shared post-treatment time trajectory, within-subject variation in circulating DN2 B cell frequency was associated with elevated plasma CXCL13 levels (β = 0.643, BH-adjusted p < 0.0001), suggesting that individual differences in CXCL13 expression explain a component of DN2 B cell abundance variation that is independent of treatment kinetics (**Supplementary Fig. 8b**).

At baseline, EF-skewed B cells additionally displayed a CXCR3^hi^ CD62^lo^ phenotype, consistent with an inflammatory tissue-trafficking profile^43^. CXCR3 surface expression was higher on DN2 than DN1 B cells before therapy (p = 0.0156, n = 7), and this phenotype occurred in the context of elevated interferon-associated chemokines. Following CAR-T cell therapy, DN2 cells contracted, CXCR3 surface expression declined and circulating interferon-associated chemokines were reduced. Although residual EF-like B cells retained a CXCR3 transcriptional imprint, this program was diminished in plasmablast/plasma cell populations (**Fig. 4f**). These findings indicate attenuation of the CXCR3-linked EF-to-PB trafficking axis after deep B cell depletion.

The T cell compartment showed concordant remodeling. Longitudinal flow cytometric analysis (full gating strategy shown in **Supplementary Fig. 9**) revealed transient T cell lymphopenia following CAR-T cell therapy, which normalized over the time of follow-up. At 6 months post therapy 9/16 (56.25%) of patients had reached normal CD3^+^ cell counts, Additionally, we observed no significant changes in the composition of CD4^+^ T cells, but a significant decrease of CD8^+^ T_EMRA_ cells at ≥12 months post therapy compared to baseline (p = 0.013; Supplementary Fig. 10).

In accordance with increased baseline CXCL13 levels, further flow cytometric analysis revealed that PD1^hi^ CD4^+^ T cells, encompassing both CXCR5^−^ Tph and CXCR5^+^ Tfh subsets, were markedly expanded in active SLE compared to healthy controls (p = 0.0003; **Fig. 4g**). Following CAR-T cell therapy, PD1^hi^ T cells declined significantly in responders and resembled healthy controls. At 12-months, Tph cells and Tfh cell frequencies had fully contracted, supporting a disruption of peripheral B-helper programs consistent with a “circuit breaker” mechanism in which deep B cell depletion disrupts reciprocal T-B cell activation (**Fig. 4h,i**).

Regulatory T cells, which were reduced at baseline consistent with SLE-associated Treg dysfunction, underwent a transient expansion in responders peaking at approximately 6 months and normalizing by 12 months (**Fig. 4j**).

Collectively, these findings demonstrate that CD19 CAR-T cell therapy creates a window of B cell aplasia, during which the inflammatory environment can collapse. In responders, this window is accompanied by a progressive normalization of the inflammatory milieu: Interferon signature resolves, CXCL13 and EF-promoting chemokines are suppressed, pathogenic PD1^hi^ Tph cells contract and regulatory T cells transiently expand. Thus, newly generated B cells are released into a remodeled milieu, which does not appear to favor EF-differentiation, as evidenced by their lack of re-entry into the aNAV-DN2-plasmablast trajectory.

#### Failed milieu reset precedes SLE relapse

CAR-T cell therapy induced substantial clinical improvement: SLEDAI-2K decreased by ≥4 points in all patients, corticosteroids were discontinued in all patients and 15 of 16 evaluable patients with a minimum 6-month follow-up achieved DORIS remission. Two patients did not achieve durable complete responses: Patient #14 showed complete response to treatment regarding LN, but exhibited hypocomplementemic vasculitis refractory to therapy (potentially representing a separate disease entity; persistent hypocomplementemia highlighted in **Extended Fig, 1**). Follow-up was ended at month 6, as further immunosuppressive treatment was initiated. Patient #9 initially achieved complete remission (SLEDAI-2K = 0) but subsequently developed fulminant relapse of SLE (**Fig. 5a**). Given the otherwise uniform response, Patient #9 constituted a rare natural experiment enabling direct interrogation of which events distinguish successful immune reset from its failure and was analyzed separately from the responder cohort after day 203.

**Fig. 5.**
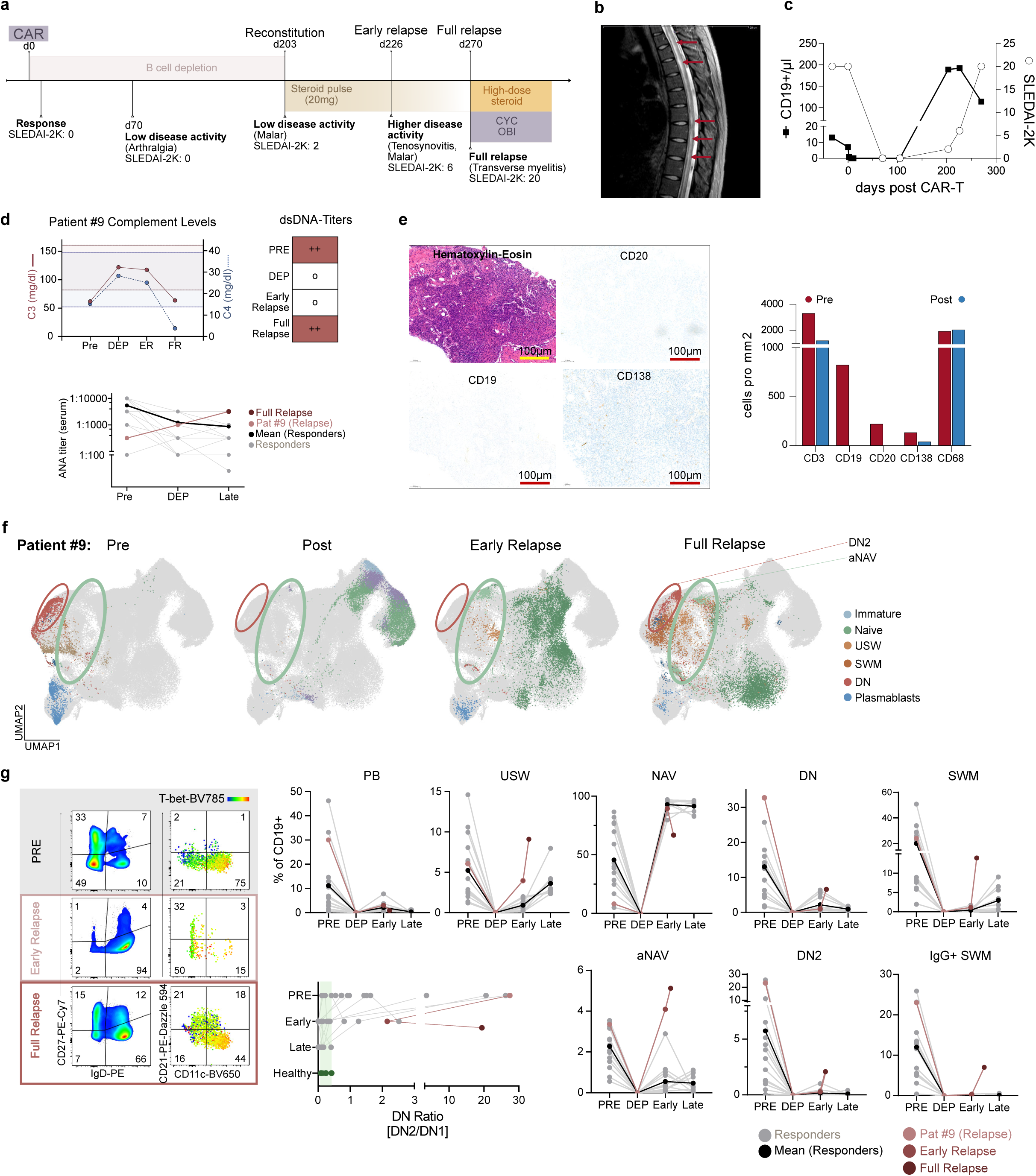
Immediate extrafollicular differentiation of reconstituting B cells precedes disease relapse. **a**, Clinical course of the relapsed patient (patient #9). Time is shown as days after therapy, with key clinical milestones annotated. The duration of B cell depletion and the timing of treatment initiation are indicated. SLEDAI-2K scores are provided at the highlighted timepoints. **b**, Spinal MRI of patient 9 at Day 270. Sagittal T2-weighted MRI of the thoracic spine. Red arrows indicate longitudinal intramedullary T2 hyperintensity spanning multiple thoracic vertebral segments, consistent with transverse myelitis. **c**, Peripheral blood B cell counts and SLEDAI-2K scores are shown over time in the relapsing patient. B cell counts are plotted on the left y-axis as CD19^+^ cells/µl derived from flow cytometry, SLEDAI-2K scores are on the right y-axis. Time is shown as days after CD19-CAR-T cell therapy. **d**, Serum C3 and C4 concentrations are shown for the relapsing patient at the indicated timepoints. Anti-dsDNA antibody levels assessed by semiquantitative analysis are shown at baseline (pre), during B cell depletion (DEP) and at the latest available follow-up (post). Connected dots represent individual patients. The relapsing patient (patient #9) is highlighted (n = 1), and the black line indicates the mean titer of treatment responders (n = 17). ANA titers from serum are shown for the responding patients and the relapsed patient #9 (bold line: mean). **e**, Representative Hematoxylin-Eosin staining, CD19, CD20 and CD138 immunohistochemical staining is shown for inguinal lymph node tissue obtained at day 70 post therapy (post) of patient #9 during peripheral B cell aplasia. Quantification of CD19^+^, CD20^+^, CD3^+^, CD138^+^ and CD68^+^ cells is presented as cells per mm² for tissue-based quantification before therapy and at d70. **f,** UMAP of B cells from flow cytometry data, highlighting cell type composition in the relapsed patient compared to all non-relapsed patients. Colors indicate B cell subsets, aNAV and DN2 B cells are highlighted by the ellipses. **g**, Representative flow cytometry plots are shown for the relapsing patient at baseline (pre), early relapse (day 226) and full relapse (day 270). Corresponding plots on the right display intracellular T-bet expression projected as a color gradient. Frequencies of the indicated B cell subtypes as percentage of CD19^+^ cells are shown for the same timepoints, with the individual relapse trajectory overlaid on the corresponding responder data, including individual longitudinal courses, as in Fig. 2b and Fig. 3h. DN ratio is shown analogous to Fig. 3h, the relapsed patient is highlighted in red.

Following initial remission, patient #9 developed arthralgia at day 70, which resulted in later arthritis and tenosynovitis. B cell reconstitution occurred at day 203 and was followed by progressive increase in disease activity (SLEDAI-2K = 6 at day 226). At day 270, the patient developed a fulminant relapse (SLEDAI-2K = 20) with severe cutaneous manifestations (**Fig. 5a**) and transverse myelitis (representative MRI shown in **Fig. 5b**). Thus, overt SLE activity coincided with, and then rapidly followed, B cell return in the peripheral blood (**Fig. 5c**). In contrast, conventional serological markers remained less informative during early relapse: complement levels and anti-dsDNA antibodies stayed within the normal range, whereas ANA titers rose only in this patient (**Fig. 5d**), establishing how conventional serological markers lacked sensitivity to detect impending reactivation of disease.

Serial inguinal lymph node biopsies were performed in this patient before treatment and during peripheral B cell aplasia at day 70 after CD19 CAR-T cell therapy. The day 70 biopsy showed depletion of CD19^+^ and CD20^+^ B cells (**Fig. 5e**), loss of follicular dendritic cell networks and absence of germinal center structures (**Supplementary Fig. 11**). These findings confirm effective tissue-level B cell depletion during aplasia and argue against persistence of organized germinal center activity as immediate source of relapse.

The trajectory of B cell reconstitution in patient #9 diverged markedly from durable responders. At first reappearance (day 203), reconstituting B cells showed a naïve phenotype, constituting mainly transitional and resting-naïve B cells by flow cytometric characterization (**Fig. 5f**). Post-therapy scRNA-seq of the relapsing patient was performed during the stage of early signs of SLE return at day 228; B cell characterization revealed a predominantly naïve reconstituting B cell compartment, accompanied by expansion of intermediate B cell states, and was analyzed in parallel with paired responder samples (**Extended Fig. 5a–c**). Consistent with de novo B cell generation after CD19 CAR-T therapy, paired BCR profiling showed a shift towards IgM/IgD heavy chain usage throughout the cohort, including exclusive detection of IgM/IgD clones after therapy in the relapsing patient (**Extended Fig. 6a**). Somatic hypermutation was concomitantly reduced across reconstituting B cell subsets, including memory and EF-like compartments in responders and relapse alike, further supporting de novo B cell development (**Extended Fig. 6c, Supplementary Fig. 12**).

Clonal BCR diversity distinguished the clinical trajectories: Responders maintained a broadly polyclonal repertoire after therapy, with BCR diversity metrics increasing post therapy without evidence of oligoclonal outgrowth. The relapsing patient, however, showed markedly reduced diversity, suggesting selective expansion of a restricted or clonally focused, newly generated B cell repertoire during clinical recurrence (**Extended Fig. 6b**, **Supplementary Fig. 13**).

Rather than persistence of this naïve phenotype that uniformly characterized the first months of reconstitution in responders, Patient #9 showed immediate and prominent expansion of CD21^lo^ CD11c^+^ aNAV B cells from day 228. This rapidly progressed to accumulation of DN2 cells less than two months later and was accompanied by IgG^+^ class-switched memory B cells and plasmablasts, hence fully recapitulating the pathological B cell architecture of active SLE (**Fig. 5f,g**). In contrast to transient aNAV appearances in some responders, this patient showed full execution of the EF differentiation cascade. Their fast return fits the rapid nature of EF B cell differentiation^27^ and suggests that the state of the systemic immune environment influences B cell differentiation fate.

IGHV4-34 usage, known to be linked with autoreactivity, was enriched after CAR-T (**Extended Fig. 6**), consistent with naïve-skewed B cell reconstitution where IGHV4-34 is physiologically represented^35^. In relapse, however, IGHV4-34 usage extended into EF-like, DN1 and activated memory compartments, indicating failure to restrain autoreactive-prone clones from effector differentiation (**Extended Fig. 6, Supplementary Fig. 14**). Thus, with IGHV4-34 positive naïve B cells appearing in remission, relapse requires both, a germline encoded autoreactive substrate and a permissive inflammatory milieu; in responders, the remodeled environment appears sufficient to prevent expansion of these potentially autoreactive clones despite their presence in the naïve pool.

Single-cell pathway analysis showed broad post-treatment attenuation of interferon-α, interferon-γ and inflammatory-response programs across B cell subsets in patients in remission, consistent with release from the inflammatory state of active SLE (**Extended Fig. 5e,f**). By contrast, relapse was marked by strong re-emergence of interferon-related programs across the entire regenerated B cell compartment, including EF-like cells (**Extended Fig. 5e,f**). Thus, the returning B cells in relapse did not merely re-acquire an EF phenotype; they did so within a transcriptionally inflammatory state that mirrored the persistent systemic interferon milieu.

Critically, in the relapsed patient the pathological immune environment preceded B cell reappearance. Unlike responders, Patient #9 exhibited persistently elevated serum CXCL13 and IFN-ω as well as memory related serum protein markers that failed to normalize during the post-therapy window of B cell aplasia (**Fig. 6a**, **Supplementary Fig. 2c**). This observation raises a mechanistic paradox: CXCL13 is canonically produced by FDCs within lymphoid structures, yet serial lymph node biopsy had confirmed complete germinal center abrogation during peripheral B cell aplasia (**Fig. 5e**).

**Fig. 6.**
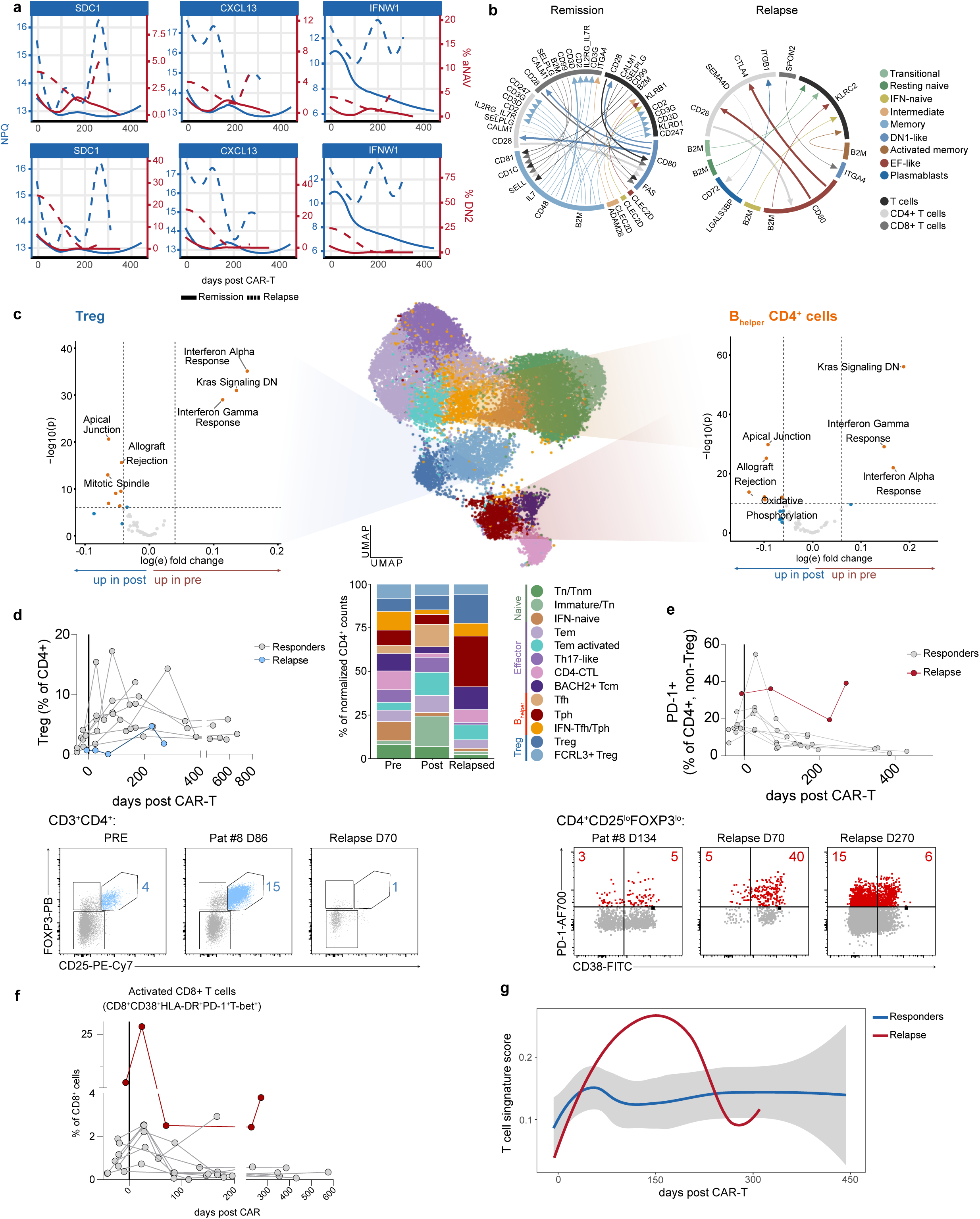
Persistent inflammatory milieu and expanded activated T cells before B cell recovery and SLE relapse. **a**, Model-derived trajectories of serum SDC1, CXCL13 and IFN-ω (IFNW) are shown for responders (blue; n = 10) alongside the trajectories of flow cytometry-derived frequencies of activated naïve B cells and DN2 cells (red). Solid lines show median in patients maintaining remission. The relapsing patient (n = 1) is shown as an individually modeled dotted line. **b**, Comparison of ligand-receptor interactions between B cells-T cells communicating during remission and relapse. Chord diagrams show the top10-ranked interactions enriched in remission and relapse, grouped by B cell subset and T cell compartment. Colors denote the sender cell subset, and line width reflects the difference in interaction ranking between conditions. **c**, UMAP shows annotated CD4⁺ T-cell populations from single-cell sequencing, with volcano plots highlighting differential Hallmark pathway activity in Treg (left) and B-helper CD4⁺ cells (right). Below, stacked bar plots show the relative distribution of CD4^+^ cell states across timepoints. **d,** Tregs were defined as CD3^+^CD4^+^CD25^+^FoxP3^+^ by flow cytometry. Individual trajectories are shown for responders and for relapse. Representative gating plots are shown for a responder at baseline and day 86, and for the relapse at day 70. **e**, Longitudinal dynamics of PD-1^+^ non-Treg among CD4^+^CD25^lo^FoxP3^−^ CD4^+^ T cells after therapy. Individual patient trajectories are shown for responders and the relapse. Representative gating shown for a responder at day 134 and for relapse at day 70 (B cell aplasia) and day 270 (full relapse). **f**, Activated CD8^+^ T cells were identified as CD3^+^CD8^+^CD38^+^HLA-DR^+^PD-1^high^ by flow cytometry. Individual patient frequencies (percentage of CD8^+^) are shown (n = 10). The relapsed patient is highlighted. **g**, Corresponding, time-delayed kinetics were seen in proteomic analysis of immune-checkpoint proteins CD274, CTLA4, ICOSLG, LAG3, PDCD1, with transient expansion kinetics seen in the relapsed patient (light blue; n = 1) compared to responders (dark blue; n = 9).

Single-cell analyses further indicated that relapse was accompanied by reactivation of B-T cell communication (**Supplementary Fig. 15**). Ligand-receptor analysis showed enrichment of inferred interactions between EF-like B cells and CD4^+^ T cells during the stage of early relapse, including CD80-dependent costimulatory/checkpoint axes and additional EF-like B cell-T cell interactions within the most enriched programs (**Fig. 6b**). The relapsed patient’s CD4^+^ cells showed re-emergence of interferon-related pathway activity together with expression of genes linked to co-stimulation and B-helper function, including *CD28*, *ICOS*, *PDCD1* and *MAF*, which were increased together with adhesion molecules such as *ITGA4* and *ITGB1*, consistent with reactivation of a tissue-interacting B-helper-like CD4^+^ T cell state (**Fig. 6c**, **Extended Fig. 7**). Consistent with renewed B cell-T cell crosstalk, relapse was accompanied by relative overrepresentation of B-helper-like CD4^+^ T cell states by single-cell analysis (**Fig. 6c**) and persistence of PD1^hi^ non-Treg CD4^+^ T cells by flow cytometry (**Fig. 6e**), positioning the persistence of B-helper T cells including Tph cells as a possible explanation for the elevated CXCL13 levels during B cell aplasia. Thus, the relapse state was not characterized by reappearance of B cells per se, but by re-engagement of a B-T cell circuit which fueled pathologic EF differentiation.

This occurred in parallel with impaired regulatory re-equilibration. Patient #9 was the only individual who failed to show the transient increase in circulating Treg frequencies observed in responders (**Fig. 6e**). Although relapse-associated CD4^+^ T cells displayed regulatory/checkpoint-associated transcripts, they showed reduced *TGFB1* expression, arguing against effective TGF-β-associated regulatory re-equilibration. This was also not accompanied by expansion of canonical Tregs by flow cytometry, suggesting an incomplete or ineffective regulatory counterbalance occurring in the relapsed patient (**Fig. 6e**, **Extended Fig. 7**). On the B cell side, relapse-associated EF-like cells retained TGF-β receptor expression (*TGFBR1/2*) but lacked coordinated induction of downstream TGF-β response, as shown by reduced expression of TGF related hallmarks (**Extended Fig. 5f**) and related genes including *SMAD3*, *KLF10*, *ID3*, *TGIF1* (**Extended Fig. 7i**). Together with reduced *TGFB1* in CD4^+^ T cells, this argues for incomplete TGF-β-mediated regulatory imprinting

In addition, activated HLA-DR^hi^ PD-1^+^ T-bet^+^ CD8^+^ T cells transiently expanded before B cell reappearance, and serum checkpoint-associated proteins, including soluble CTLA4, PDCD1, CD274, LAG3 and ICOSLG, showed stable kinetics or slightly declined over the first 12 months post therapy in serum of responders, but an early rebound during aplasia in the relapsing patient rather than the progressive decline observed in responders (**Fig. 6f,g**).

Collectively, these findings suggest a model in which SLE relapse resulted from a coordinated failure of immune re-equilibration. In this model, PD1^hi^ T cells and the CXCL13/interferon milieu persisted through B cell aplasia, the homeostatic Treg expansion failed, and nascent B cells were immediately channeled into the pathogenic EF trajectory upon reconstitution via reactivation of EF-like B cell–T cell communication, followed by clinical relapse. The reappearance of aNAV and DN2 cells, alongside rising CXCL13, preceded clinical relapse by weeks, identifying these as potential sentinel signals for impending relapse.

## Discussion

Beyond documenting clinical responses, this study provides one of the first mechanistic accounts of how deep B cell depletion translates into durable immunological remission in SLE. Our data indicate that durable drug-free remission following CD19-CAR-T cell therapy in SLE requires not merely deep B cell depletion extending to tissue, but the lasting elimination of the extrafollicular B cell differentiation trajectory. It further establishes that the window of B cell aplasia is itself a dynamic immunological event, during which the inflammatory milieu must normalize to ensure durable remission. We propose a feed-forward loop model in which the interferon-rich milieu and PD1^hi^ T peripheral helper cells may provide the signals (IFN-λ, IL-21, CXCL13) that drive naïve B cells through the trajectory from aNAV to DN2 to plasmablasts, generating autoantibody-secreting cells that amplify tissue inflammation and perpetuate the cycle.

Underlying this cascade is the view that instead of being intrinsically pathogenic, B cells in SLE appear to be environmentally corrupted and shaped into a pathogenic state by chronic exposure to interferons, CXCL13 and T peripheral helper cell-derived signals. CAR-T cell therapy breaks this loop at the B cell node. Deep tissue-level depletion eliminates not only circulating B cells, but also B cells embedded in tissue niches, as confirmed by serial lymph node biopsies and consistent with prior reports^37,38^.

Given the high response rate of CAR-T cell therapy in SLE, the relapse (while limited to one case) constituted a natural experiment that offered a rare opportunity to interrogate the conditions for failure of immune reset. Despite comparable depth of B cell depletion, as confirmed by prolonged aplasia and complete B cell depletion in the lymph node, the inflammatory milieu persisted: CXCL13 and IFN-ω remained elevated during B cell aplasia, PD1^hi^ CD4^+^ T cells failed to contract and the homeostatic Treg expansion did not occur. Reconstituting naïve B cells hence entered a functionally pathogenic environment: We propose that they were immediately channeled into the extrafollicular trajectory, culminating in DN2 expansion and fulminant relapse. These results are in line with SLE relapse post CAR-T cell therapy being driven by newly-generated B cells rather than treatment-resistant clones in murine SLE models^46^.

Crucially, lymph node biopsies confirmed complete absence of follicular dendritic cell networks and T-follicular helper cells post therapy, suggesting that class-switched differentiation during relapse occurred through the extrafollicular pathway. This observation satisfies the criteria recently proposed by Eisenbarth et al.^47^ for demonstrating extrafollicular responses and suggests that the extrafollicular pathway alone may be sufficient to generate the full pathogenic B cell repertoire in SLE. This rapid emergence of class-switched cells is consistent with TLR- and cytokine-driven activation in the EF compartment, supported by prior evidence that BAFF, APRIL and type I interferons can license class-switch outside the GC^19,20^. The persistence of elevated CXCL13 in the absence of follicular dendritic cells implies an alternative cellular source, consistent with the expanded PD1^hi^ T cell population that remained elevated throughout B cell aplasia. This finding aligns with accumulating evidence positioning Tph cells as principal providers of extrafollicular B cell help through IL-21 and CXCL-13 secretion in non-lymphoid tissues including the kidney^22^ and extends it by demonstrating that Tph persistence may sustain the pathogenic niche even after complete B cell elimination. The contraction of Tph cells in responders as compared to relapse, supports a circuit-breaker mechanism in which deep B cell depletion disrupts reciprocal T-B cell activation, though direct tissue-level evidence for this source was not obtained.

The contrast with rituximab is instructive. Anti-CD20 therapy only transiently decreases extrafollicular intermediates including DN2 cells, and these effects abrogate within months, with DN repopulation predicting treatment failure^48,49^. In contrast to rituximab, CAR-T cell therapy achieves B cell depletion in tissue niches, resulting in durable suppression of extrafollicular subsets. The qualitative difference is not the magnitude of peripheral B cell reduction but the capacity to disrupt tissue-resident extrafollicular niches sufficiently to trigger environmental remodeling. The permanent disappearance of anti-dsDNA autoantibodies in all patients implicates CD19^+^ short-lived plasmablasts sustained by ongoing EF differentiation, rather than long-lived plasma cells, as their predominant cellular source, with important implications for why deep CD19 depletion succeeds where anti-CD20 therapy often fails. The slow, progressive normalization of interferons and CXCL13 over months supports gradual tissue remodeling secondary to sustained absence of B cell-T cell crosstalk. A key effector cytokine in this circuit, IL-21, was not represented in the NuLISA panel and was not reliably detected at the transcript level in our scRNA-seq data; longitudinal quantification of IL-21 therefore remains an important next step to directly confirm Tph-mediated EF B cell instruction in this setting.

Further supporting this reset model, IgM⁻ naïve B cells, consistent with chronic BCR engagement and downregulation^42^, were lost after therapy. BCR signaling was re-established across B cell subsets during reconstitution; this suggests that B cells in SLE are not intrinsically locked into a pathogenic state, but are shaped by ongoing chronic stimulatory cues, particularly as IgM^lo^ naïve B cells subsets have been implied in the generation of EF-like B cell subsets^50,51^.

These findings anchor the concept of immune reset, which is defined functionally as sustained remission but still lacks a mechanistic definition (reviewed by Junt et al.^36^ and Schett et al.^52^). We demonstrate that immune reset is not a single event but a cascade which begins with depletion, propagates through environmental remodeling and is sustained by reconstitution of naïve B cells into a milieu no longer permissive for pathogenic differentiation. Importantly, the observation that reconstituting B cells in responders followed healthy ontogeny argues against models exclusively invoking intrinsic B cell defects, such as genetic variants in TLR7 signaling or epigenetic imprinting as the sole driver of autoimmunity; rather, the environment dictates whether nascent B cells become pathogenic. Notably, this concept may not extend to monogenic forms of SLE, which manifest early in life and where these homeostatic networks are likely to be overridden^11,32,53^. However, given the uniform pattern seen across responders spanning diverse manifestations and treatment histories, our observations support the primacy of the environment over B cell-intrinsic programming as the determinant of pathogenic differentiation in polygenic disease context. As B cell-depleting approaches are moving towards precision-targeting^54^, understanding how B cell-intrinsic autoreactive potential intersects with a permissive immune milieu will be essential to define immune reset, predict therapeutic durability and guide patient stratification.

Several clinical implications follow. First, DN2 cells and serum CXCL13 may serve as sentinel biomarkers for extrafollicular pathway re-engagement, detecting impending relapse weeks before conventional markers such as complement consumption and anti-dsDNA titers. Second, patients in whom the inflammatory milieu fails to normalize during B cell aplasia, as identifiable by persistent CXCL13 or interferon elevation, may benefit from adjunctive therapies such as type I interferon blockade, BAFF inhibition or Tph-targeted approaches deployed during the window between depletion and reconstitution. Third, the observation that the reconstituting compartment remained overwhelmingly naïve for more than three years has implications for vaccination strategy and infection risk management.

This study has important limitations. First, the cohort comprises patients from a single center, at young age and predominantly European background. Second, an intriguing mechanistic insight into SLE relapse derives from a single patient. More relapses after CAR-T cell therapy will need to be studied. Third, while lymph node biopsies provide tissue-level validation, which is not yet available in other studies, further tissue-level validation of extrafollicular pathway truncation in responders will be important. In the long run, interventional studies, such as adjunctive interferon blockade in patients with persistent CXCL13 during B cell aplasia, will be needed to further clinically validate this model. Environmental factors, including recently implicated viral contributors such as Epstein-Barr virus^55^, were not captured in this analysis. Clarifying their course during long-term follow-up will be essential to define their role in sustained remission vs. relapse.

Nevertheless, and looking forward, this framework moves the field from a depletion-centric view of B cell therapy towards one centered on the quality of immune reconstitution and the permissiveness of the systemic immune environment and thus provides a mechanistic foundation for the rational design and monitoring of tolerance-restoring therapies in autoimmunity.

## Methods

### Patients and treatment

Eighteen patients with SLE were included, all fulfilling the 2019 EULAR/ACR classification criteria. Disease activity was assessed by rheumatologic evaluation and SLEDAI-2K scoring. All patients underwent CD19-CAR-T cell therapy at the Department of Medicine 3 (Rheumatology and Immunology), University Hospital Erlangen, after interdisciplinary evaluation by a specialized board of rheumatologists and hemato-oncologists. Patients were either enrolled in the CASTLE trial (NCT06347718; EudraCT: 2022-001366-35; EUCT: 2024-516819-24-00) or treated under compassionate-use approval.

Thirteen of 18 patients (72%) were female, with a median age of 24 years (IQR, 20.75-38.75) and a median disease duration of 5 years (IQR, 2-15). At treatment initiation, all patients had active SLE, with a median SLEDAI-2K score of 15 (range, 6-22), indicating moderate-to-high disease activity. Active skin involvement was present in all patients. Fifteen of 18 patients (83%) had biopsy-confirmed class II, III, or IV lupus nephritis, of whom 9 of 15 (60%) had active renal involvement at inclusion, defined by active urine sediment, macroproteinuria, or hematuria (Supplementary Fig. 1d). Detailed patient characteristics and leading organ manifestations are provided in **Table 1**. Before CAR-T cell therapy, patients had received a median of 6 prior therapies (IQR, 4.75-7.25), and 9 of 18 (50%) had previously been treated with a B cell-depleting anti-CD20 monoclonal antibody (rituximab or obinutuzumab). Two patients were in B cell aplasia at the time of CAR-T cell treatment owing to prior rituximab exposure yet still showed high disease activity (SLEDAI-2K 20 and 22, respectively).

All patients received standard lymphodepletion with one cycle of fludarabine and cyclophosphamide, followed by a single infusion of the advanced therapy medicinal product Zorpocabtagene Autoleucel (MB-CART19.1) containing autologous CD19-CAR-transduced CD4^+^/CD8^+^-enriched T cells generated from peripheral blood apheresis products using a lentiviral vector, as previously described^1,39^.

### Sampling time points and follow-up

Peripheral blood samples were collected longitudinally from all 18 patients at multiple follow-up timepoints after CAR-T cell therapy. To harmonize analyses across patients despite interindividual differences in B cell reconstitution kinetics, four key immunological reference timepoints were defined: **Pre** (baseline during active disease), **Depletion** (DEP; post-therapy during B cell aplasia), **Early** (first timepoint of B cell reappearance), and **Late** (stable B cell reconstitution at ≥12 months). For the late timepoint, the most recent available sample was used for each patient. In this subgroup, the median follow-up was 24 months (IQR, 14-29.25) after B cell reappearance and 30 months (IQR, 19.75-32.5) after CAR-T cell therapy. Follow-up details and sampling intervals are provided in **Supplementary Table 1**, and a schematic overview of longitudinal B cell follow-up is shown in **Extended Fig. 1**. Subset frequencies were compared with healthy control donors. Each represented value indicates a separate sample and measurement. Additionally, peripheral blood of age- and sex-matched healthy controls was drawn; the characteristics of the donors are shown in **Supplementary Table 2**.

### Ethics

Written informed consent was obtained from all participants before sample collection in accordance with the Declaration of Helsinki. The use of patient samples and clinical data was approved by the institutional review board of the University Hospital Erlangen (license 334_18B). Healthy donor samples were collected after written informed consent.

### PBMC isolation, flow cytometry and flow analysis

PBMC were isolated using SepMate-50 (Stemcell Technologies) and Ficoll-Paque gradient centrifugation (1200 x g for 10 minutes at room temperature) according to manufacturer’s protocol. Absolute cell counts were measured using BD Trucount tubes (BD Biosciences) according to the manufacturer’s instructions.

The detailed staining protocol for B and T cell characterization is shown in **Supplementary Table 3**. Fixable Viability Stain 780 (Thermo Fisher Scientific) was included for live/dead discrimination. Samples were acquired on a spectral flow cytometer (Cytek Nothern Lights (Fremont)) or an LSRFortessa (BD Biosciences (Germany)) and analyzed using the FlowJo software (v.10; TreeStar). B cells were identified as live singlet CD19^+^ lymphocytes. Within the B cell compartment, immature/transitional, naïve, memory, and plasmablast populations were defined based on CD20, CD27, CD38, CD24, CD21, and immunoglobulin isotype expression. Memory B cells were further separated into pre-switched and class-switched subsets according to IgD, IgM, IgG, and IgA expression. In addition, SLE-associated activated or extrafollicular-like B cell populations were assessed using CD11c, T-bet, CD21, HLA-DR, CD72, and CXCR5. Identification of B cell subsets and atypical B cell compartment was analyzed in accordance with the more recently published guidelines for a uniform cytometric B cell analysis^56^ and the recently defined roadmap for EF-response definition ^47^. A representative gating strategy are shown in **Supplementary Fig. 1**.

For T cell analysis, live singlet CD3^+^ lymphocytes were separated into CD4^+^ and CD8^+^ T cell populations. Naïve and memory T cell subsets were defined using CD45RA, CD45RO, CD27, and CD62L expression. Activated T cells were assessed based on CD38 and HLA-DR, and PD-1 was used to evaluate activated or exhaustion-associated phenotypes. Regulatory T cells were identified within the CD4^+^ compartment as CD25^+^FOXP3^+^ cells, while T helper polarization was further assessed by T-bet expression. Representative gating strategies are shown in **Supplementary Fig. 9**. All measurements were acquired from individual patient samples.

High-dimensional flow cytometry data from 18 patients and five healthy donors were analyzed in R (version 4.5.2). Automated quality control was performed using flowAI (v1.40.0) package^57^. Marker intensities were arcsinh-transformed using sample- and channel-specific cofactors. Processed files were imported as flowSet objects and converted into Seurat (v5.4.0) v5 objects. Dimensionality reduction was performed using UMAP via uwot (v0.2.4), graph-based clustering was performed using Leiden clustering, and technical variation was corrected using Harmony (v1.2.4). Clusters were manually annotated based on canonical B cell marker-expression patterns.

Differential abundance was tested using patient-level cell counts. For major B cell compartments and sufficiently represented subtypes, counts were modeled using negative binomial mixed models with time point as fixed effect, sample identity as random intercept, and total B cell count as offset (n_cells ∼ timepoint + (1 | sample), offset by log(total). Rare subtypes with near-complete absence across time points were modeled using binomial mixed models (cbind(n_cells, total - n_cells) ∼ timepoint + (1 | sample)). Model diagnostics were assessed using simulated residuals from DHARMa (v0.4.7), and pairwise contrasts were estimated with emmeans (v2.0.2) using Bonferroni correction^58,59^.

Differential marker-expression analysis was performed at the patient level. For each B cell subtype, marker expression was summarized per sample and time point. Linear mixed-effects models were fitted using lme4 (v2.0.1), with marker expression modeled as a function of time point, marker identity and their interaction, and sample identity included as a random intercept (value ∼ timepoint * marker + (1 | sample)). Marker-wise, pairwise contrasts between time points were estimated using emmeans with Bonferroni correction^60^. A summary of included patient samples is shown in **Supplementary Table 4**.

### Autoantibody quantification

Autoantibodies against dsDNA were detected by commercially available ELISA (Orgentec; Catalog #: ORG 507; cutoff: 40 U ml−1).

### Immunohistochemistry

Inguinal lymph node immunohistochemistry was performed as recently published ^38^. Staining was performed on 2-µm-thick FFPE tissue sections on positively charged adhesive slides (TOMO) using routine protocols on the VENTANA BenchMark ULTRA platform (Ventana). Following deparaffinization and CC1-based antigen retrieval, sections were stained as described above; representative CD19 and CD20 stainings are shown in **Figure 5**. Slides were scanned on a Hamamatsu S210 at x400 magnification and analyzed in QuPath (v0.4.3). Regions of interest were annotated and analyzed by a pathologist, and CD19-, CD20-, CD68- and CD138-positive cells were quantified per mm^2^,

### Single-cell sequencing data acquisition and preprocessing

Single-cell transcriptomic and immune repertoire data were analyzed from 15 samples, including 10 in-house Erlangen samples from patients undergoing CD19 CAR-T therapy and five publicly available samples from PRJNA1303131 from patients treated with BCMA/CD19 CAR-T therapy using R (version 4.5.2). Sample availability from both cohorts is shown in **Supplementary Table 4**. FASTQ files were processed using Cell Ranger (v10.0, 10x Genomics) with the cellranger multi pipeline and default parameters. Gene expression reads were aligned to the 10x Genomics human GRCh38 2024-A reference (refdata-gex-GRCh38-2024-A). V(D)J reads were aligned to the 10x Genomics human V(D)J GRCh38 reference (refdata-cellranger-vdj-GRCh38-alts-ensembl-7.1.0).

The Erlangen cohort comprised previously published in-house samples from PRJNA1100403 and PRJNA1100404 together with three newly generated hashtag-multiplexed samples. The three newly generated hashtag-multiplexed samples were prepared using the 10x Genomics Chromium Single Cell 5′ Gene Expression v3 chemistry with paired-end sequencing, following the manufacturer’s protocol. Two samples (#12 and #17) had paired pre and post treatment time points, whereas sample #9 was available only at the post time point.

### Empty droplet removal

Empty droplets were removed using the emptyDrops function from DropletUtils (v1.30.0) with test.ambient = TRUE, niters = 15000, and lower = 50. Barcodes with an FDR ≤ 0.001 were retained for downstream analysis^61^.

### Ambient RNA correction

Ambient RNA contamination was corrected using SoupX (v1.6.2). Preliminary clusters required for contamination estimation were generated from the filtered expression matrix using a standard Seurat (v5.4.0) workflow, including normalization, variable feature selection, scaling, PCA, nearest-neighbor graph construction using 30 principal components, and Leiden clustering. Contamination fractions were estimated using autoEstCont with forceAccept = TRUE, and corrected integer count matrices were generated using adjustCounts with roundToInt = TRUE^62^.

### Cell annotation

Reference-based cell type annotation was performed using SingleR (v2.12.0) with the celldex Monaco Immune Data reference. Both fine-grained (label.fine) and broad (label.main) immune-cell labels were generated. These automated annotations were then manually curated using canonical marker expression and clustering structure to define the final cell type labels used for downstream analyses^63^. The top 20 differentially expressed genes in the annotated B and T cell clusters and shown in **Supplementary Table 5, 6**.

### Cell-level quality control

Cells were retained if they satisfied all the following criteria: mitochondrial gene fraction < 10%, number of detected genes > 200 and < 5,000, log10 genes per UMI > 0.8, and singlet classification by scDblFinder (v1.24.10).

### Data integration, dimensionality reduction, and clustering

Following quality control, Seurat assay layers were joined using JoinLayers. Gene expression matrices were normalized using NormalizeData, and variable features were identified using FindVariableFeatures. Variability from V(D)J genes was suppressed using scRepertoire quietVDJgenes (v2.6.2) before downstream dimensionality reduction. Data were scaled with ScaleData, and principal component analysis was performed using RunPCA. Batch correction and integration were performed using Harmony^64^.

Nearest-neighbor graph construction and UMAP embedding were performed using the Harmony reduction with FindNeighbors and RunUMAP, respectively. Cell clusters were identified using Leiden clustering with FindClusters.

### Pseudo-bulk differential expression

For pseudobulk differential expression analysis, raw RNA counts were aggregated separately for each annotated cell type by subject and visit using Seurat AggregateExpression. Analyses were performed separately for CART19-treated samples, BCMA CART19-treated samples and the combined dataset. Only subjects with paired pre and post samples were retained. The relapsed patient #9 was also removed. Differential expression was tested with DESeq2^65^ using the design ∼ patient + condition, where patient accounts for paired sampling and condition represents post versus pre. Log_2_ fold changes were shrunk using lfcShrink with type = “ashr”. The genes included when calculating composite module score are shown in **Supplementary Table 7,8**.

### Gene Set enrichment analysis

Pre-ranked gene set enrichment analysis was performed separately for B cell and T cell subsets using fgsea. For each cell type, genes were ranked by the DESeq2 Wald statistic. Hallmark, Reactome and Gene Ontology Biological Process gene sets were obtained from MSigDB using msigdbr. Gene sets were tested separately for each collection, and pathways with Benjamini–Hochberg-adjusted P < 0.05 were considered significant.

### Pseudotime and trajectory inference

Pseudotime was inferred using Palantir through SeuratExtend package on the Harmony-integrated embedding ^66,67^. Transitional B cells from PRE samples were set as the start state, and POST plasmablasts/plasma cells were set as the terminal state. For trajectory-based differential expression, pseudotime was recalculated after excluding the relapsed patient #9 and Memory/Activated memory B cells. Generalized additive models were fitted using tradeSeq (v1.24.0) fitGAM on highly variable genes, with separate smoothers for each visit-treatment condition^68^. The resulting tradeSeq genes associated with B cells across the pseudotime for different treatment conditions are shown in **Supplementary Table 9**.

### Repertoire analysis

BCR and TCR contigs were processed using the Immcantation framework^69^. Contig FASTA files generated by Cell Ranger were aligned to germline V(D)J reference sequences using IgBLAST through AssignGenes.py. AIRR-formatted rearrangement tables were then generated using MakeDb.py igblast together with the corresponding 10x Genomics contig annotation files and IMGT germline reference sequences. Extended annotation was enabled during database generation.

BCR rearrangements were imported from AIRR-formatted Immcantation output and matched to B cell scRNA-seq metadata using reformatted cell barcodes. Productive rearrangements with consistent heavy-, kappa- or lambda-chain annotations were retained. Cells with more than one productive heavy-chain rearrangement were excluded to avoid ambiguous BCR assignment.

TCR rearrangements were imported in AIRR format and matched to annotated T cell scRNA-seq metadata. Productive in-frame TRA and TRB rearrangements without stop codons were retained, requiring non-missing CDR3 amino-acid sequences, V and J gene calls, and UMI count ≥ 2. For each cell, the dominant TRA and TRB rearrangement was selected by highest UMI count. TRB clonotypes were defined as subject-specific combinations of TRBV gene, TRB CDR3 amino-acid sequence and TRBJ gene.

### Cell-cell communication analysis

Cell-cell communication analysis was performed using the LIANA R package (v0.1.12), a ligand–receptor inference framework for single-cell RNA-seq data. Analyses were run on annotated cell populations using default LIANA settings. Putative ligand–receptor interactions were inferred from cell type–level expression profiles and ranked using LIANA-derived interaction scores. Selected interactions were visualized for biologically relevant sender–receiver cell type pairs.

### Proteomics platform and preprocessing

Circulating protein concentrations were measured using the NuLISA targeted proximity ligation immunoassay platform (Alamar Biosciences), which quantifies 248 analytes from a single low-volume plasma sample. Relative quantification (NPX-equivalent, log₂ scale) values were used throughout. A focused panel of B cell-relevant proteins and interferon-pathway analytes (N = ∼30 proteins) was selected for all pharmacodynamic analyses, which were performed using R (version 4.5.2). Subjects without a pre-treatment sample and samples appearing as outliers in the first two principal components were omitted from analyses. The final pharmacodynamic dataset comprised n = 11 SLE subjects with samples collected from day −46 to day +514 relative to CAR-T infusion. Nine of the 11 pre-treatment samples were collected the week before infusion. A healthy control (HC) cohort of 10 samples was included for reference.

### Pharmacodynamic modeling: subgroup trajectory analysis

To identify proteins with differential post-treatment trajectories according to baseline clinical subgroups, normalized protein abundance (NPX) was modeled as a function of post-treatment time, clinical subgroup, and the time x subgroup interaction, per analyte. Time was modeled with a natural cubic spline basis with four degrees of freedom, capturing both linear and nonlinear temporal trends. Intra-subject correlation across repeated measurements was accounted for by estimating a consensus within-subject correlation using limma (blocking on subject ID). Model fitting and empirical Bayes moderation of test statistics were performed using the limma package^70^. F-statistics were computed on the joint set of time × subgroup interaction coefficients. P-values were adjusted for multiple comparisons across all proteins using the Benjamini-Hochberg false discovery rate (FDR) method.

### Comparison to healthy controls

To quantify normalization of protein levels towards the healthy state following B cell depletion, NPX values for SLE patients at each timepoint were expressed as Z-scores relative to the HC distribution, with a robust MAD-based SD estimate and a minimum SD floor of 0.2. For visualization, per-protein Z-scores from all SLE subjects were interpolated onto a common 50-point time grid using LOESS and displayed as a heatmap.

To formally test whether the post-treatment protein trajectory of SLE patients differed from the HC baseline, a limma-based model was fitted to the combined SLE + HC dataset. The design included an SLE/HC indicator and an SLE-specific natural spline time term (nsSLE), so that the spline captures only within-SLE temporal variation relative to the (time-invariant) HC mean. An F-test on the joint nsSLE spline coefficients identified proteins with significant trajectory deviation from HC levels; a t-test on the groupSLE intercept term identified proteins with significant baseline elevation at the time of infusion. Both tests used empirical Bayes moderation (robust trend) and BH-adjusted p-values.

### Baseline associations with clinical metadata

Spearman rank correlation coefficients between baseline protein levels and continuous clinical variables (e.g., B cell subset frequencies, disease duration, age) were computed for all protein–variable pairs. Mann–Whitney U tests were used to compare baseline protein levels between the two levels of each binary clinical variable. Baseline protein levels in SLE patients were compared to HC values using a similar test, followed by BH correction across all proteins. Effect sizes were computed as the difference in group means (NPX scale).

### Association of cell abundance with proteomics

The rmcorr package^71^ was used to compute repeated-measures Pearson correlations between longitudinal proteomics measurements and contemporaneous B cell subset frequencies by flow cytometry (B cell subsets: plasmablasts, DN, SWM, IgG⁺ SWM, USW, NAV, aNAV, PB). To assess co-variation between circulating protein levels and B cell subset frequencies independently of the shared treatment trajectory, linear mixed models were fitted with protein NPX as the outcome, natural cubic splines in time as a covariate, the log₂-transformed B cell subset frequency as the variable of interest, and a random subject intercept. Coefficients on the flow term were taken as estimates of time-adjusted co-variation. Models were fitted using lmerTest with maximum likelihood estimation, and p-values were BH-adjusted across all protein × flow-subset combinations. Analyses were restricted to subjects with at least two matched proteomics and flow cytometry timepoints, excluding timepoints with zero B cells.

### Survival analysis

Time-to-hypogammaglobulinemia (IgG < 610 mg/dL or IgM < 35 mg/dL) and time-to-CD19 B cell reappearance were modeled using univariable Cox proportional hazards regression, with each baseline protein and flow cytometric variable tested independently. Kaplan-Meier curves were stratified by baseline clinical subgroups. All Cox models used the baseline (pre-infusion) sample and baseline flow cytometry values as predictors. BH-adjusted p-values were computed across all tested biomarkers.

### Statistical analysis of flow cytometric data

For analysis of flow cytometric data, all statistical analyses were performed using GraphPad Prism version 10. Multiple groups from the same analysis were compared by Kruskal-Wallis test and unmatched samples were compared using non-parametric Mann Whitney Test. Dunn’s correction was applied for multiple comparisons. Indicated p values reflect ∗ p < 0.05, ∗∗ p < 0.01, ∗∗∗ p < 0.001∗∗∗∗, p < 0.0001. Error bars show median values and IQR for numeric parameter or mean values with SD for cellular frequencies as indicated in the figures.

## Data availability

All depicted data show raw data and each value represents an individual patient value. Source data for each figure is provided with this paper. Patient data is shown in a pseudonymized form through numbering. Data can be obtained from the corresponding author upon request. Single cell RNA sequencing data will be deposited in the Gene Expression Omnibus (GEO) database prior to publication.

## Code availability

No custom code was generated for this study. All analyses were performed using publicly available software and R (v4.5.2) packages as described in the Methods section. The code is available from the corresponding author upon request.

## Author Contributions

D.M.N. and K.A. designed and performed experiments, analyzed data and wrote the first draft of the manuscript. P.G.G. contributed to bioinformatics analysis and data interpretation. T.R. and L.B., contributed to flow cytometry data acquisition. F.I. contributed to bioinformatics analysis of proteomics data. N.F. and S.L.N.C. contributed to data analysis and interpretation. M.H., A.W., J.B., C.T., S.B. and R.G.B. recruited patients, collected clinical samples and curated clinical metadata. S.K., M.A. and S.V. performed CAR-T cell manufacturing and monitoring. L.M. performed serum autoantibody measurements. F.M. and A.M. supervised clinical CAR-T cell treatment for the hematology department. M.E. performed histopathological analyses of lymph node biopsies. M.G.R. and C.T. performed lymph node biopsies. A.B. contributed expertise and analytical input. G.S. initiated the CASTLE CAR-T cell trial, acquired funding and contributed to manuscript editing. R.G.B. designed the study, acquired funding, supervised the work, analyzed data, wrote the first draft of the manuscript and revised the manuscript. All authors discussed the results and approved the final version of the manuscript.

## Extended figure legends

**Extended Fig. 1.**
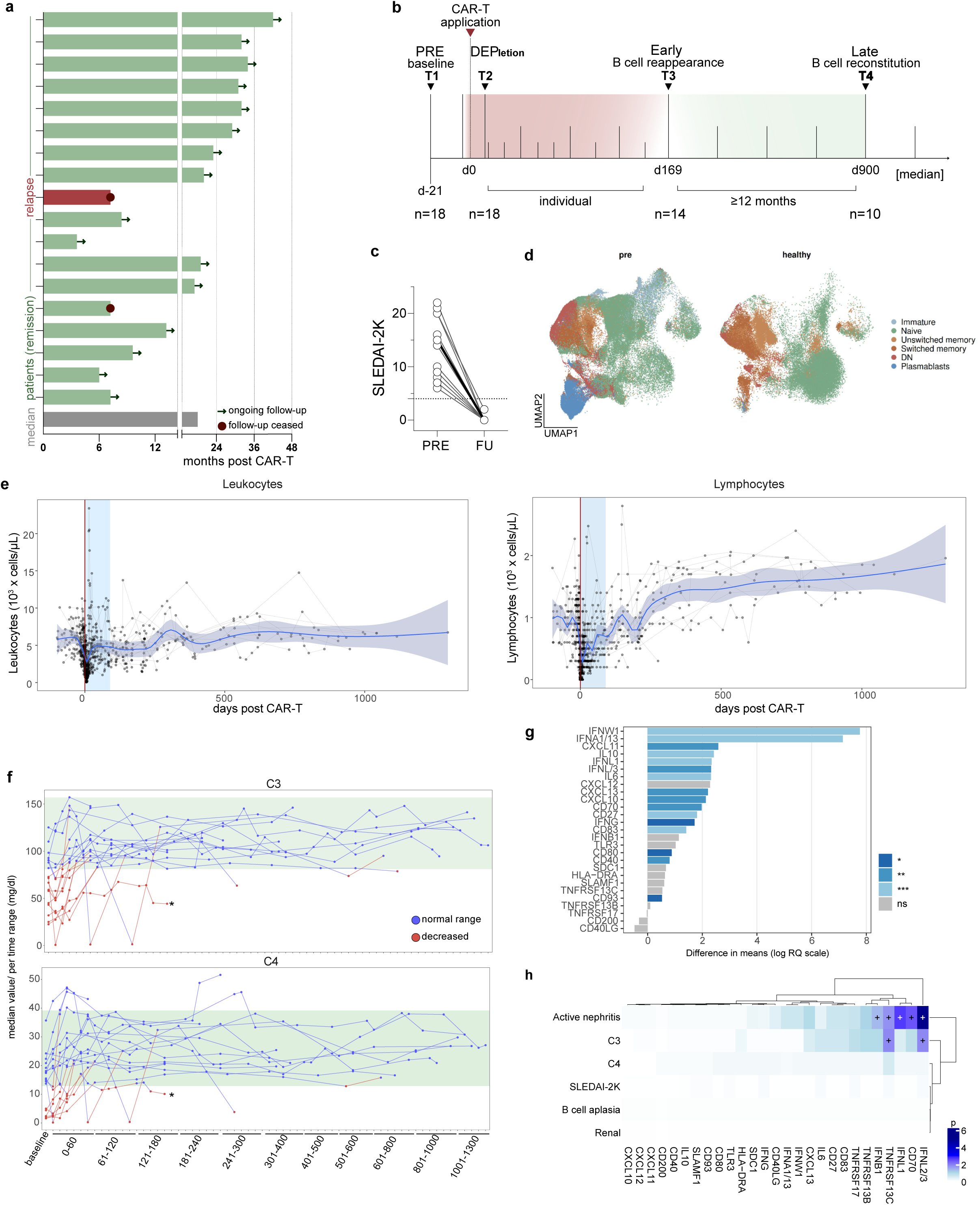
Laboratory follow-up and changes following CAR-T cell therapy. **a**, Duration of individual follow-up after CAR-T cell infusion in patients with SLE. Bars indicate months post CAR-T cell therapy; green bars indicate patients in drug-free remission and red bars indicate the relapsed patient. Arrows denote ongoing follow-up and dots indicate closed follow-up. The grey bar indicates the median follow-up duration. **b**, Schematic overview of the longitudinal sampling strategy. Samples were collected at baseline before CAR-T cell therapy, during B cell depletion, at early B cell reappearance and at late B cell reconstitution. B cell reappearance was defined as CD19^+^ B cells ≥5 cells/µl and B cell reconstitution as CD19^+^ B cells ≥60 cells/µl. **c,** SLEDAI-2K scores at baseline and latest follow-up. Lines connect paired samples from individual patients. Dotted line indicates low disease activity threshold (SLEDAI-2K = 4). **d**, UMAP of main B cell types from flow cytometry data, pre (n = 15) and healthy (n = 5) state. **e,** Longitudinal laboratory measurements after CAR-T therapy. Leukocyte and lymphocyte counts were plotted over time relative to CAR-T infusion. Red vertical line indicates CAR-T infusion at day 0. Blue shaded area marks the early post-treatment period from day 1 to day 90. Blue curves show GAM-smoothed trends with 95% confidence intervals. **f,** Longitudinal C3 and C4 complement levels after CAR-T therapy. Green area indicates the normal range. Blue points are within the normal range; red points are below the normal range. The asterisks highlight patient #14, suffering from hypocomplementemic vasculitis. **g,** Baseline serum proteomic differences between patients with SLE before CAR-T cell therapy and healthy controls. Bars indicate differences in mean protein abundance on a log relative-quantification scale. Significance was assessed by two-sided Mann-Whitney U test followed by Benjamini-Hochberg correction for multiple testing (from n = 10). Color indicates significance level. **h**, Longitudinal comparison of trajectory of serum proteins, split by indicated metadata. Statistics: Colors indicate the β-coefficient for the B cell subset term; crosses indicate associations significant after Benjamini-Hochberg correction (p ≤ 0.05).

**Extended Fig. 2.**
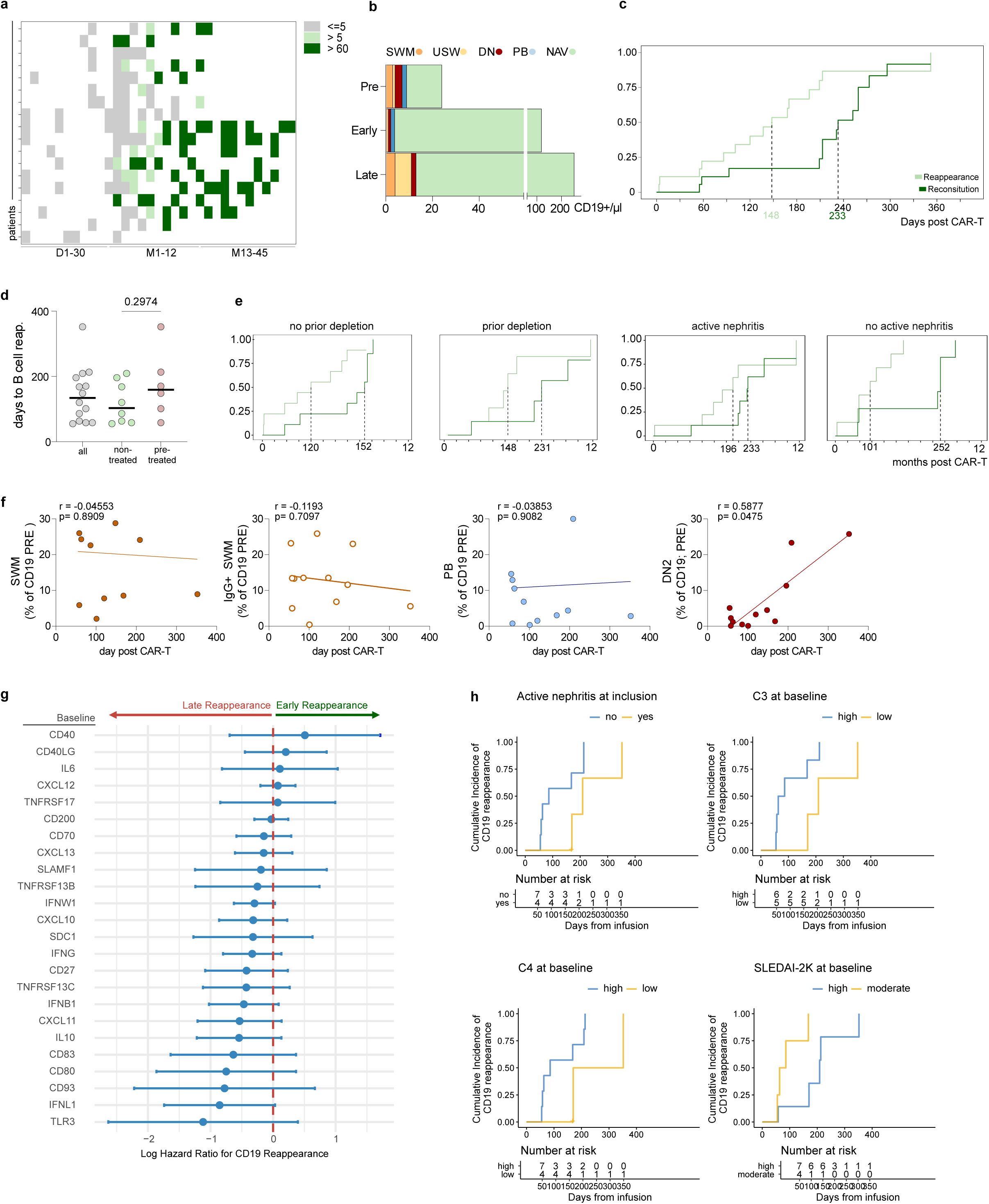
Reconstitution kinetics after CAR-T are influenced by baseline characteristics. **a,** Heatmap of peripheral B cell recovery states across follow-up after CD19 CAR-T cell therapy. Each row represents one patient, and each tile represents the B cell count at a follow-up interval. Gray indicates B cell aplasia, defined as <5 CD19^+^ B cells per μl; light green indicates B cell reappearance, defined as ≥5 CD19^+^ B cells per μl; dark green indicates B cell reconstitution, defined as ≥60 CD19^+^ B cells per μl. **b,** Absolute counts of circulating B cell subsets at baseline, early B cell reappearance and late B cell reconstitution. Bars show the contribution of switched memory B cells, unswitched memory B cells, double-negative B cells, plasmablasts and naïve B cells to the total CD19^+^ B cell count. **c,** Cumulative incidence of peripheral B cell reappearance and B cell reconstitution after CD19 CAR-T cell therapy. Reappearance was defined as ≥5 CD19^+^ B cells per μl and reconstitution as ≥60 CD19^+^ B cells per μl. Dashed vertical lines indicate median time to reappearance and reconstitution. **d,** Time to B cell reappearance according to prior B cell-depleting therapy. Dots indicate individual patients and horizontal bars indicate the median. Groups were compared using a two-sided Mann-Whitney U test. **e,** Cumulative incidence of B cell reappearance and reconstitution stratified by prior B cell-depleting therapy and active nephritis at baseline. Curves are shown as in **c**. **f,** Associations between baseline B cell subset composition and time to B cell reappearance. Scatter plots show baseline frequencies of the indicated B cell subsets among CD19^+^ B cells plotted against days to B cell reappearance after CAR-T cell therapy. Lines indicate linear regression fits. Spearman correlation coefficients and p values are shown in each plot. **g,** Baseline serum proteins associated with time to peripheral B cell reappearance. Forest plot showing log hazard ratios from univariable Cox proportional hazards models. Points indicate log hazard ratios and horizontal lines indicate 95% confidence intervals. The dashed vertical line marks a log hazard ratio of 0. Positive values indicate association with earlier B cell reappearance, whereas negative values indicate association with delayed B cell reappearance. **h,** Time to CD19^+^ B cell reappearance stratified by baseline clinical characteristics, including active nephritis, baseline C3, baseline C4 and SLEDAI-2K. Curves show the cumulative incidence of B cell reappearance; numbers below each plot indicate patients at risk over time.

**Extended Fig. 3.**
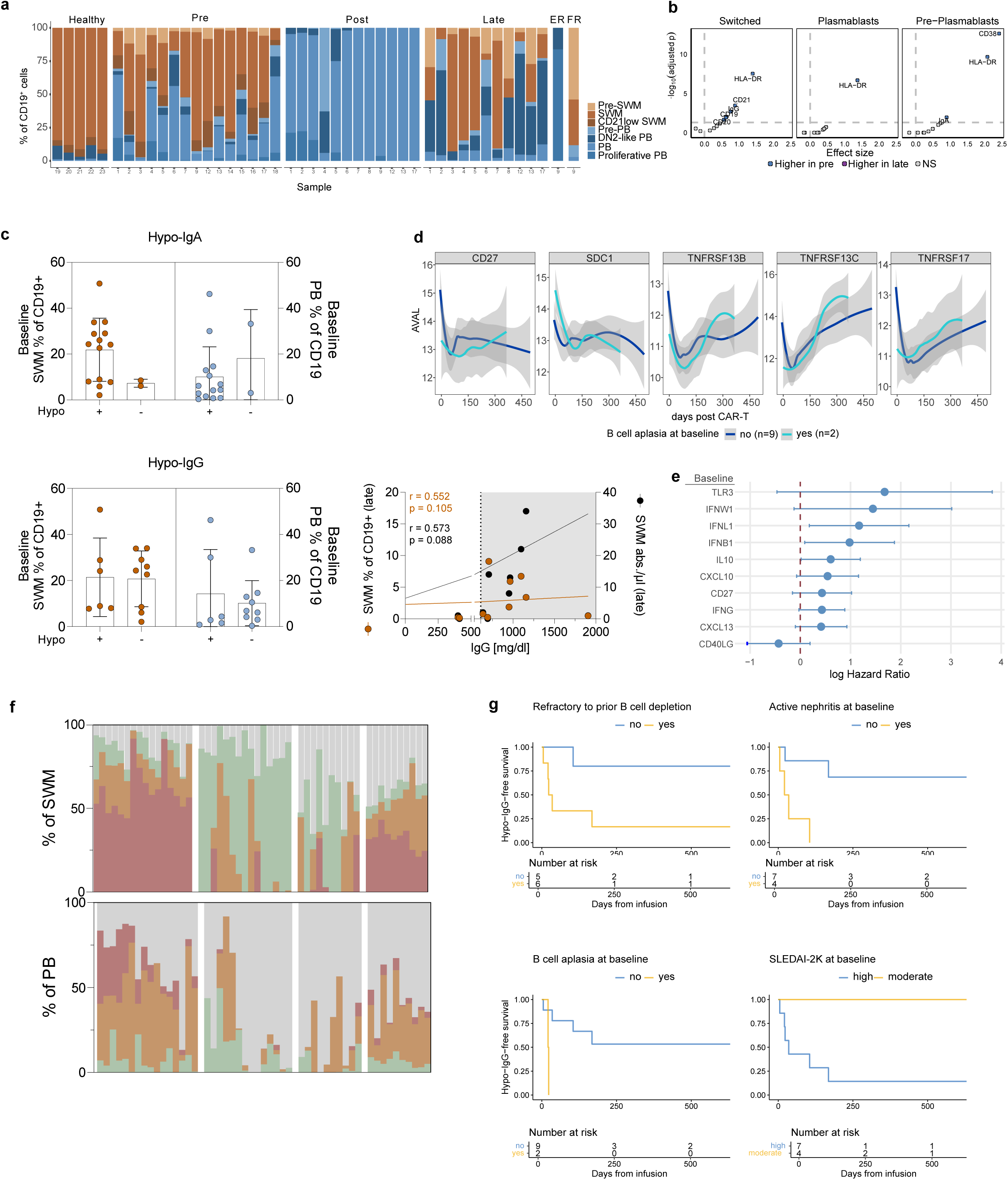
Aplasia and Hypogammaglobulinemia after CAR-T cell therapy. **a**, Distribution plot illustrates relative phenotypic shifts and cluster-specific changes in the memory and plasmablast cells during reconstitution derived from flow cytometry data. ER – early relapse, FR – follow-up relapse time point from one patient. **b,** Differential marker expression between pre-treatment and late follow-up samples within selected B cell compartments. Volcano plots show marker-level effect sizes for switched memory B cells, plasmablasts and pre-plasmablasts. Positive effect sizes indicate higher expression before treatment (blue) compared to late follow-up. P values were calculated using linear mixed-effects models with patient as a random intercept, followed by pairwise contrasts and Bonferroni correction. **c,** Baseline B cell composition according to subsequent development of hypogammaglobulinemia. Baseline frequencies of switched memory B cells and plasmablasts among CD19^+^ B cells are shown in patients with or without hypogammaglobulinemia regarding IgG or IgA after CAR-T cell therapy. The scatter plot shows the association between serum IgG levels and late switched memory B cell frequencies or absolute switched memory B cell counts. Spearman r and p values from linear regression are shown in the figure. **d,** Smoothed trajectories are shown for selected B cell activation, survival and plasma cell–associated proteins, including CD27, TNFRSF13B, TNFRSF13C, TNFRSF17 and SDC1. Lines indicate mean protein abundance over time after CAR-T cell infusion, and shaded areas indicate confidence intervals. Groups are stratified according to the presence (n = 2) or absence (n = 9) of peripheral B cell aplasia at therapy initiation. **e,** Forest plot shows log hazard ratios from univariable Cox proportional hazards models for the top 10 baseline proteins associated with development of hypogammaglobulinemia. Points indicate log hazard ratios and horizontal lines indicate 95% confidence intervals. The dashed vertical line marks a log hazard ratio of 0. **f,** Longitudinal composition of switched memory B cell and plasmablast compartments after CAR-T cell therapy. Each stacked bar represents one sample. Colors indicate the relative contribution of the indicated surface isotypes within the switched memory B cell compartment or plasmablast compartment, quantified by flow cytometric surface staining. **g,** Hypo-IgG-free survival after CAR-T cell therapy stratified by baseline clinical characteristics. Kaplan-Meier curves show time to development of hypo-IgG according to response to prior B cell depletion, presence of active nephritis at baseline, B cell aplasia at baseline and baseline SLEDAI-2K category. Numbers below each plot indicate patients at risk over time.

**Extended Fig. 4.**
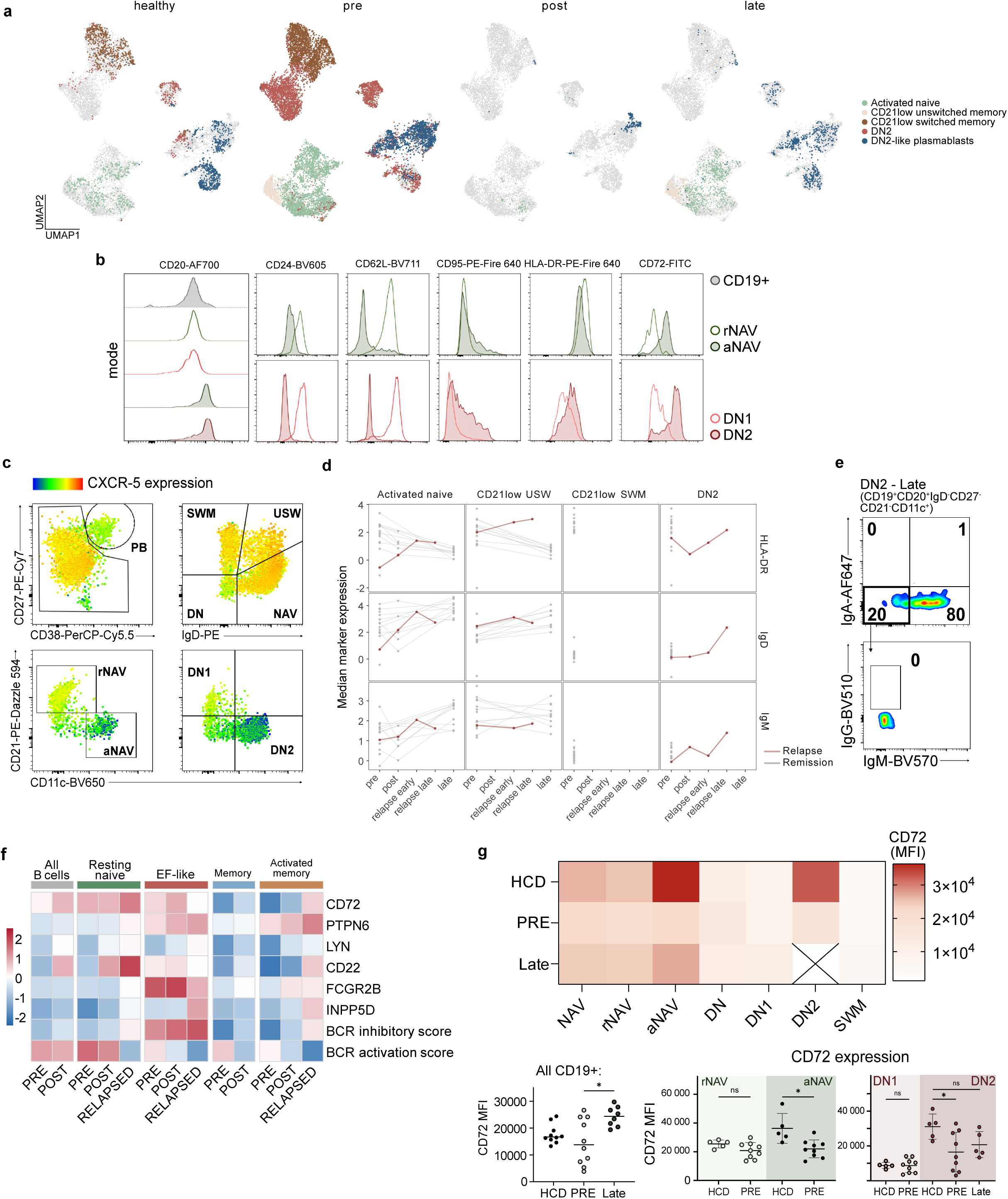
EF B cells express distinct, activation-associated features. **a**, UMAP profiling of atypical B cells subsets (CD21^lo^ CD11c^hi^ features as shown in Fig. 1b) based on flow cytometry. **b**, Representative histograms showing marker expression across CD19^+^ B cells, resting naïve and activated naïve B cells, and DN1 and DN2 cells of indicated markers. Histograms are representative of n = 5-10 individual stainings per timepoint. **c**, Representative flow cytometry plots showing CXCR5 expression across major B cell gates and CD21/CD11c-defined naïve and DN subsets. **d**, Longitudinal B cell marker dynamics by clinical outcome. Median HLA-DR, IgD, and IgM expression is shown across B cell subsets and timepoints. Red lines indicate relapsed patients; grey lines indicate patients in remission. **e**, Surface immunoglobulin expression in DN2 cells at late timepoint, showing IgA, IgG and IgM distribution, from n = 2 samples with reliably quantifiable DN2 cell count at stated timepoint. **f**, Heatmap representation of selected BCR regulatory genes and BCR pathway scores across B cell subsets and clinical stages. Colors show scaled median expression. **g,** Flow cytometric CD72 expression across B cell subsets in healthy controls, baseline active SLE and late reconstitution. Heatmap shows CD72 mean fluorescence intensity across the indicated subsets. Quantification shows CD72 expression in total CD19^+^ B cells, resting naïve and activated naïve B cells, and DN1 and DN2 cells. Statistics: Kruskal-Wallis test with post hoc Dunn‘s multiple comparison test within sample groups.

**Extended Fig. 5.**
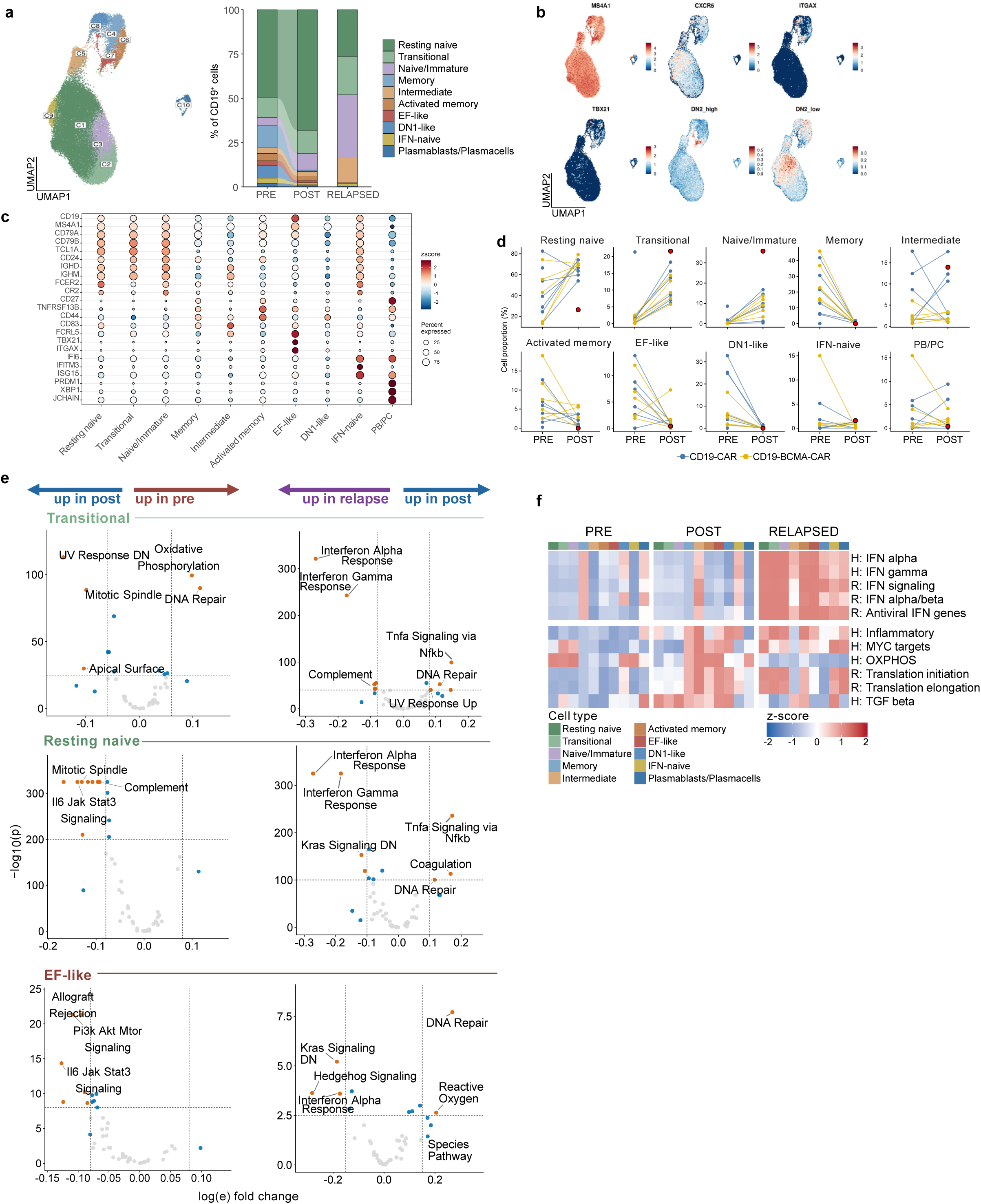
Single-cell B cell states after CAR T-cell therapy and at relapse. **a,** UMAP of annotated B cell states and their relative composition (barplots) across before (pre) and after (post) CAR-T cell therapy from scRNA data; the relapsed patient 9 is highlighted separately (relapsed). **b,** UMAP feature plots showing expression of selected marker genes and DN2 signature scores across the B cell compartment. **c,** Сanonical marker-gene expression used for B cell state annotation. **d,** Paired pre–post changes in annotated B cell state proportions from scRNA-seq data. Relapsed patient is highlighted in red. **e,** Volcano plots showing differences in AUCell pathway scores between clinical time points in transitional, resting naïve and EF-like B cells. **f,** Heatmap of selected Hallmark and Reactome pathway based on AUCell scores across annotated B cell states in pre, post samples and relapsed patient. Scores were averaged per B cell state and clinical status and converted to z-scores to visualize relative pathway activity across interferon, inflammatory, metabolic, translational and TGF-β-related programs.

**Extended Fig. 6.**
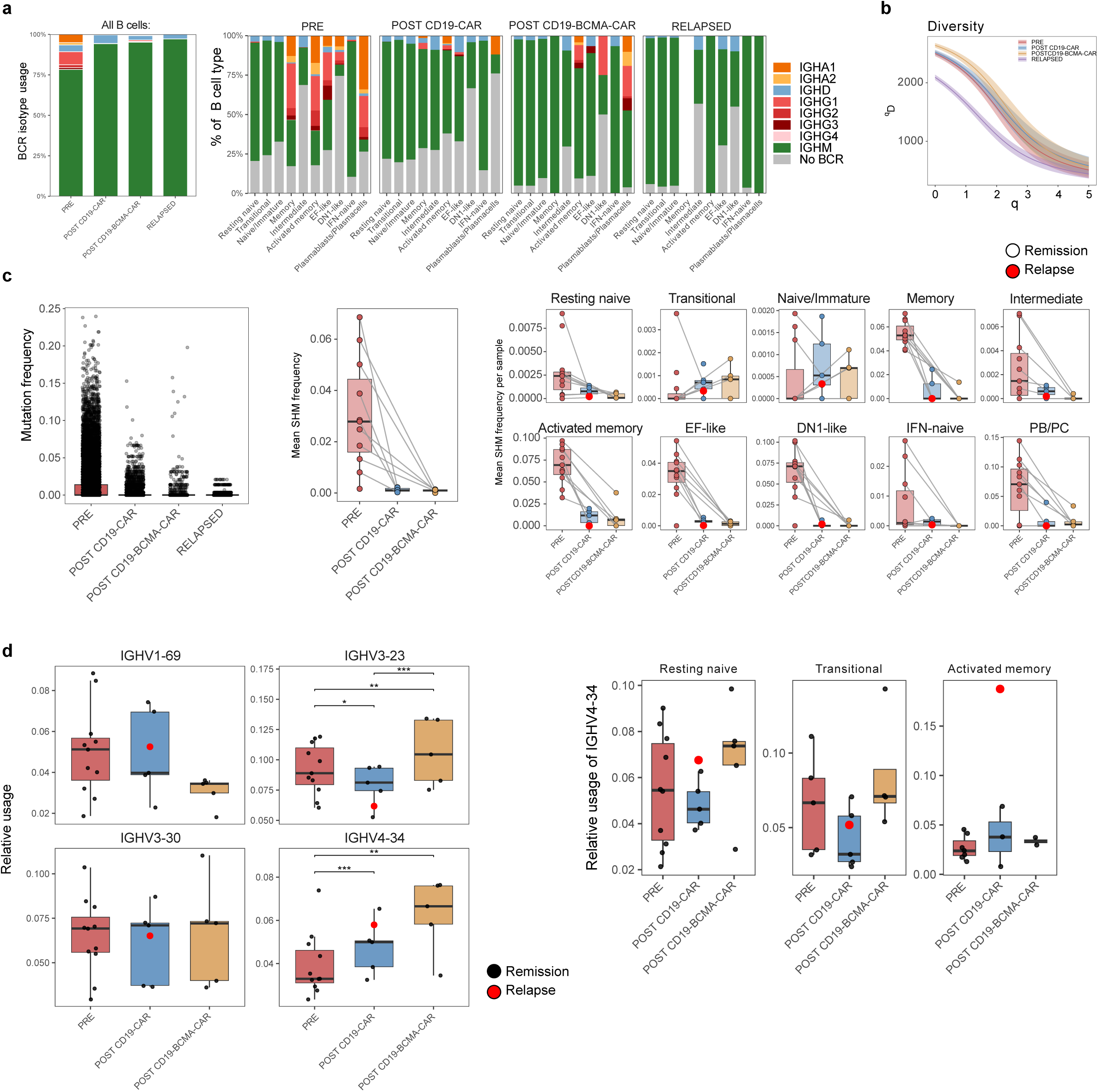
B cell reconstitution after CAR T-cell therapy is marked by an IgM-biased, low-mutational BCR repertoire. **a,** Relative immunoglobulin heavy-chain isotype usage across annotated B cell subsets at baseline (pre), after CD19 CAR-T cell therapy, after CD19/BCMA dual CAR-T cell therapy and in relapse patient. **b,** BCR repertoire diversity across clinical stages. Hill diversity curves show clonal diversity across diversity orders (q) in pre, post, and relapse sample. Shaded areas indicate confidence intervals. **c,** Somatic hypermutation across clinical stages. Left, per-cell BCR mutation frequency. Middle, mean SHM frequency per sample. Right, mean SHM frequency per sample across annotated B cell subsets. Red points indicate relapsed patient. **d**, Selected IGHV gene usage across clinical stages. Box plots show sample-level relative usage of IGHV1-69, IGHV3-23, IGHV3-30, and IGHV4-34 at pre, post CD19-CAR, and post CD19/BCMA-CAR. Red points indicate relapsed patient. **e**, IGHV4-34 usage by B cell subset. Box plots show relative IGHV4-34 usage in selected B cell subsets across the same clinical stages. Red points indicate the relapsed patient.

**Extended Fig. 7.**
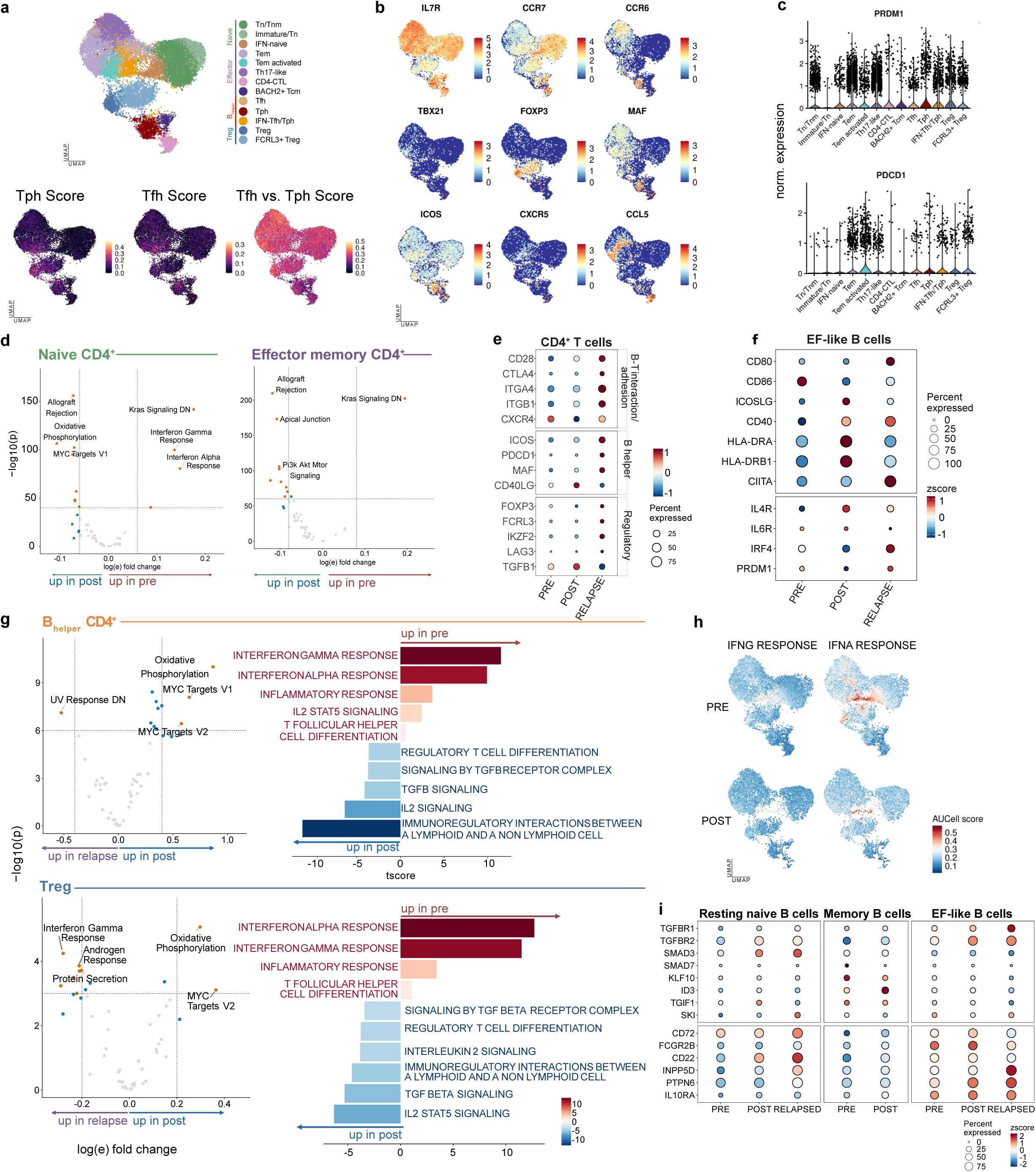
CD4^+^ T cell pathway remodeling and B cell regulatory interaction programs. **a**, UMAP of annotated CD4^+^ T cell states from scRNA-seq data (as shown in Fig. 6). Bottom panels show UCell-derived Tph, Tfh and Tfh-versus-Tph signature scores projected onto the same embedding. **b,** UMAP feature plots showing expression of selected CD4^+^ T cell marker genes, including naïve/memory-associated markers IL7R and CCR7, Th17-associated CCR6, Th1/cytotoxic-associated TBX21 and CCL5, Treg-associated FOXP3, and B helper/Tfh/Tph-associated MAF, ICOS and CXCR5. **c,** Violin plots showing expression of PRDM1 and PDCD1 across annotated CD4^+^ T cell states. **d,** Differential Hallmark pathway activity based on AUCell results between baseline and post-treatment samples in naïve and effector-memory CD4^+^ T cell families. **e,** Dot plot showing CD4^+^ T cell expression of genes linked to inferred B-T cell communication, grouped into B-T interaction, B-helper and regulatory/checkpoint programs. **f,** Differential Hallmark pathway activity in B-helper-like CD4^+^ T cells and Treg cells comparing post-treatment and relapse samples. **g,** Pathway changes in B-helper CD4⁺ and Treg cells. Volcano plots show differential pathway activity across clinical stages. Positive and negative directions indicate the clinical stage with higher pathway activity, as shown by arrows. Bar plots highlight selected pathways increased before treatment or after treatment/relapse, including interferon, inflammatory, TGF-β, IL-2, and immunoregulatory programs. **h,** UMAP projections of interferon-γ and interferon-α response based on AUCell scores in CD4^+^ T cells at baseline and after treatment. **i,** Dot plot showing expression of regulatory cytokine-response and inhibitory checkpoint-associated genes in resting naïve, memory and EF-like B cells across baseline, post-treatment and relapse samples.

