## Supplementary Figures for "A Feed-Forward Loop Between Extrafollicular B Cell Differentiation and the Inflammatory Milieu Governs Remission and Relapse in Systemic Lupus Erythematosus"

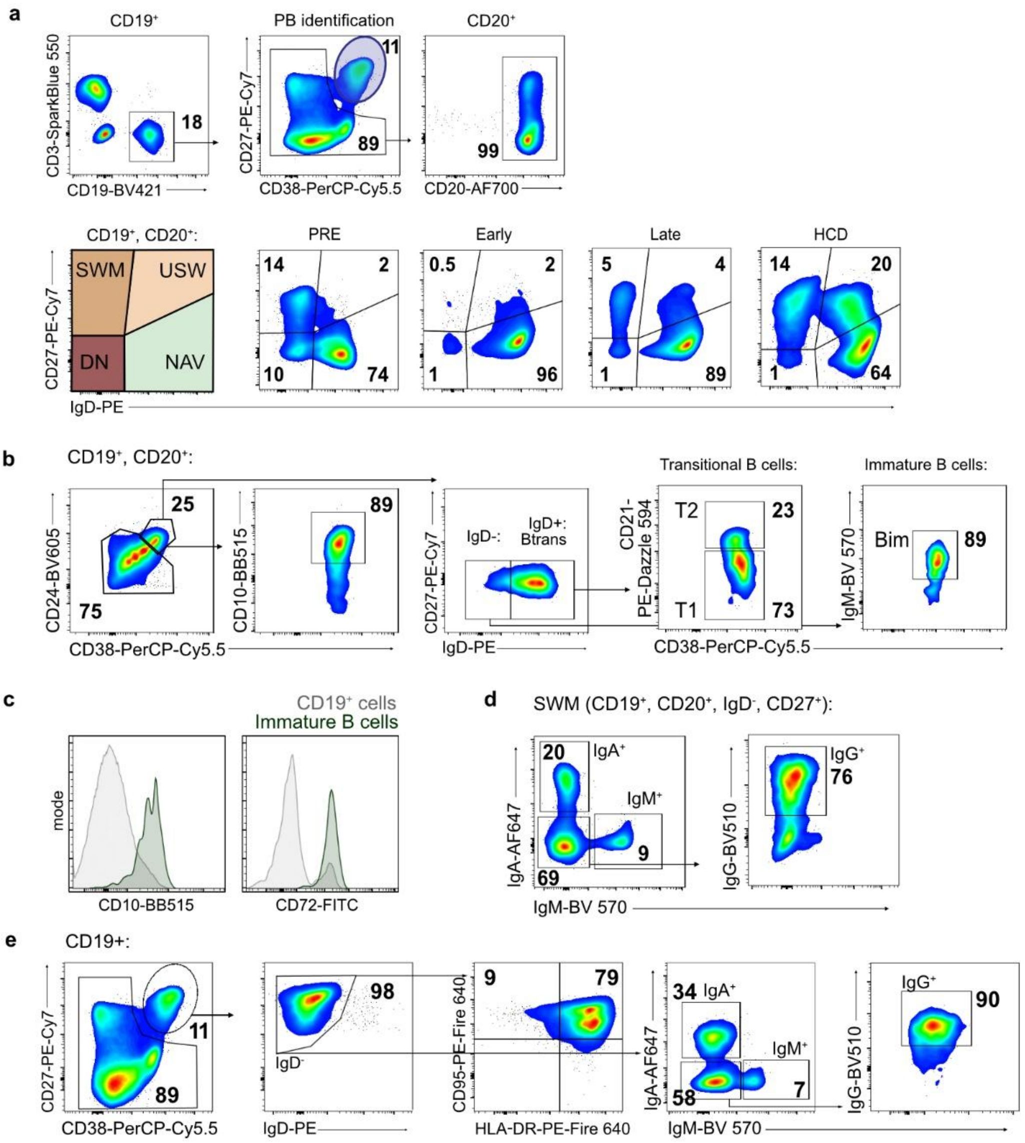


### Supplementary Fig. 1 │ Gating strategy for B cell subsets

Representative flow plots showing gating strategy to identify B cell subsets. Peripheral blood PBMC were stained with antibodies against live/dead fixable dye and doublets were excluded (not shown). **a**, Within CD3$^-$CD19$^+$ cells, plasmablasts were identified as CD27$^{++}$CD38$^{++}$. Other B cells were further identified as CD20$^+$. Within those, NAV (IgD$^+$, CD27$^-$), USW (IgD$^+$, CD27$^+$), SWM (IgD$^-$, CD27$^+$), DN B cells (IgD$^-$, CD27$^-$) are shown. **b**, Immature B cell subsets were identified within CD19+CD20+ cells and identified as CD24$^{++}$CD38$^{++}$CD10$^+$. Herein, IgD$^-$ IgM$^{hi}$ immature B cells and IgD$^+$ transitional B cells are shown. Within the latter, transitional type 1 (T1; CD21$^{lo}$) and transitional type 2 (T2; CD21$^{hi}$) are gated. **c**, Representative histograms are shown for the expression of CD10 and CD72 on immature B cells compared to overall CD19$^+$ cells. **d**, Further, within gated SWM, cells were subdivided by surface immunoglobulin isotype. IgA$^+$ and IgM$^+$ cells were identified in the first plot, while IgG$^+$ cells were defined within the IgA$^-$IgM$^-$ fraction. **e**, Plasmablasts were gated within CD19$^+$ B cells as CD27$^{++}$CD38$^{++}$ cells and further refined as IgD$^-$; additionally, HLA-DR$^{high}$/l$^{ow}$ fractions are shown. Cells were subsequently subdivided by surface immunoglobulin isotype into IgA$^+$, IgM$^+$ and IgG$^+$ fractions.

Bold numbers indicate gate frequencies from representative plots. NAV, naive B cells; USW, unswitched memory B cells; SWM, switched memory B cells; DN, double negative B cells; PB, plasmablasts; T1, type 1 transitional B cells; T2, type 2 transitional B cells; Bim, immature B cells.

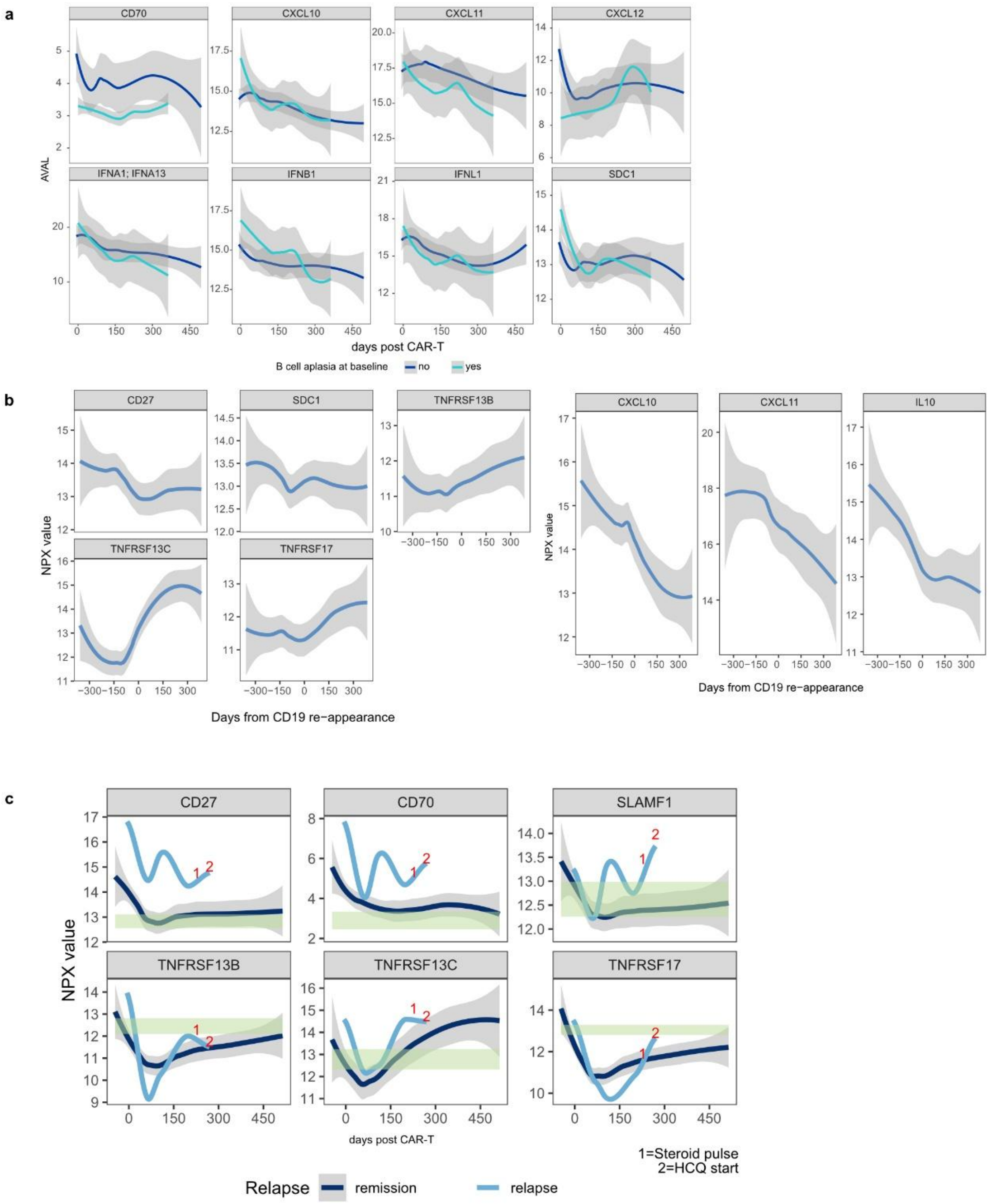


### Supplementary Fig. 2 | Serum proteomic trajectories stratified by active nephritis and relapse

**a**, Longitudinal serum proteomic trajectories after CD19 CAR-T cell therapy stratified by active nephritis at inclusion (from n = 9 with active nephritis and n = 2 without active nephritis). Shown are selected inflammatory, interferon-associated and B cell activation proteins. Lines indicate smoothed protein trajectories in patients with active nephritis and without active nephritis; shaded areas indicate confidence intervals. **b**, Serum proteomic trajectories aligned to peripheral $CD19^{+}$ B cell reappearance. The x axis shows days relative to $CD19^{+}$ B cell reappearance, with day 0 indicating the first detection of B-cell return. Shown are selected B cell activation, survival and plasma-cell-associated proteins. **c**, Longitudinal serum proteomic trajectories in patients maintaining remission (n = 10) compared with the relapsing patient (n = 1). Shown are selected B cell activation, survival and plasma cell-associated proteins, including CD27, CD70, SLAMF1, TNFRSF13B, TNFRSF13C and TNFRSF17. Normalized protein abundance (NPX) is shown as values over time after infusion. The green shaded area indicates the healthy-control reference range. Red numbers indicate clinical interventions in the relapsing patient: 1, steroid pulse; 2, hydroxychloroquine start. NPX, normalized protein expression. AVAL, analysis value.

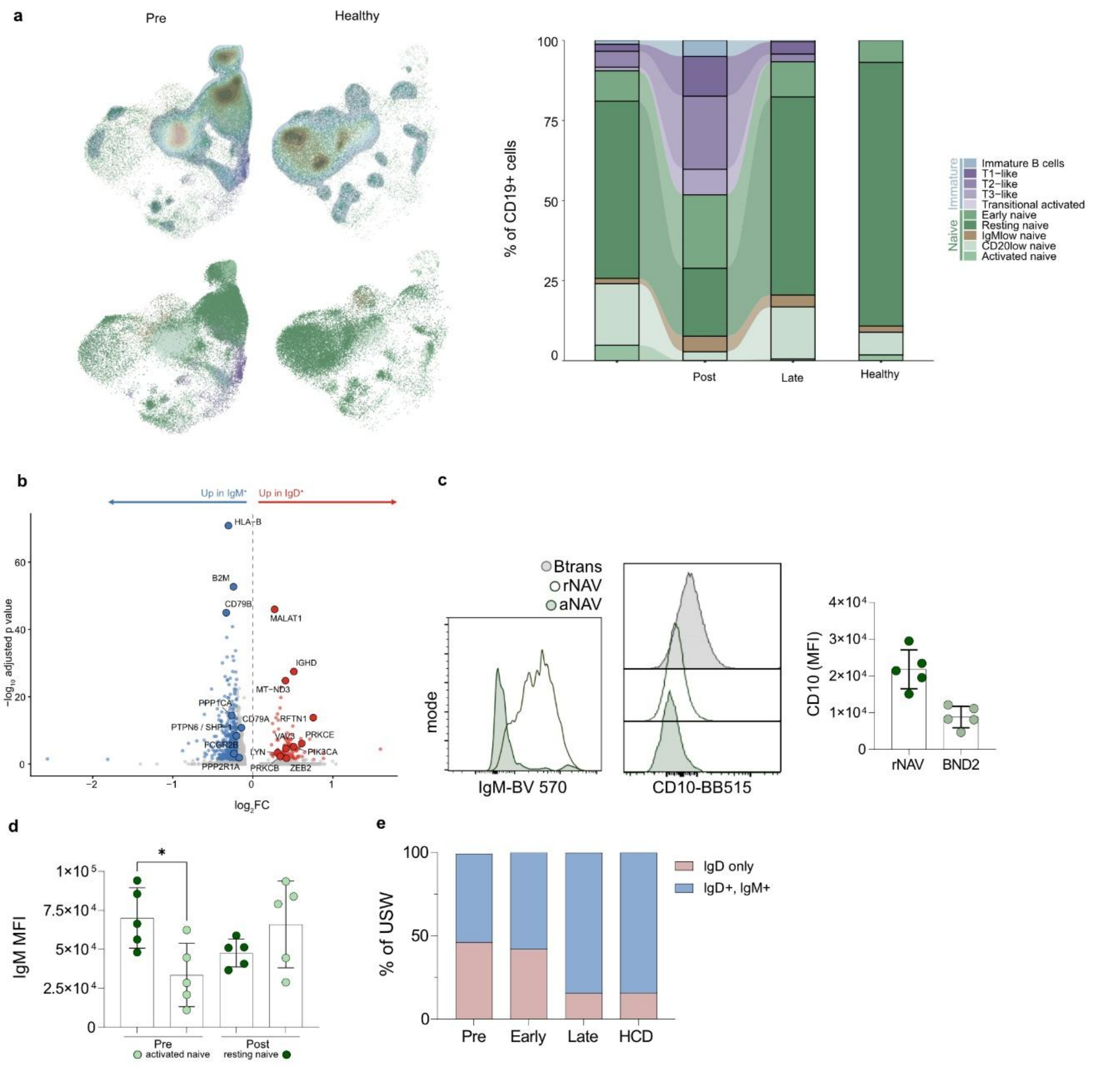


### Supplementary Fig. 3 | Naive B cell subset remodeling and IgM-low/IgD-only phenotypes after CD19 CAR-T cell therapy

**a**, UMAP and barplot of flow cytometry-defined naive B cell subsets in patients with SLE before and after CD19 CAR-T cell therapy (n = 16) and in healthy controls (n = 5). Subsets include immature, transitional-like, transitional activated, early naive, resting naive, $IgM^{low}$ naive, $CD20^{low}$ naive and activated naive B cells. **b**, Differential gene expression between $IgM^{+}$ and $IgD^{only}$ naive B cells, as identified by BCR sequencing. Volcano plot shows $\log_2$ fold changes and adjusted p values; genes enriched in $IgM^{+}$ cells are shown on the left and genes enriched in $IgD^{only}$ cells on the right. **c**, Phenotypic comparison of transitional B cells, resting naive B cells and activated naive/BND2-like cells. Representative histograms show IgM and CD10 expression; quantification shows CD10 mean fluorescence intensity in resting naive and $B_{ND2}$ cells. **d**, IgM expression in activated and resting naive B cells before and after CAR-T cell therapy. Bars indicate median values with individual samples shown as points. Groups were compared as indicated. **e**, Relative contribution of $IgD^{only}$ and $IgD^{+}IgM^{+}$ cells within the unswitched memory B cell compartment across baseline, early reappearance, late reconstitution and healthy controls. Btrans, transitional B cells.

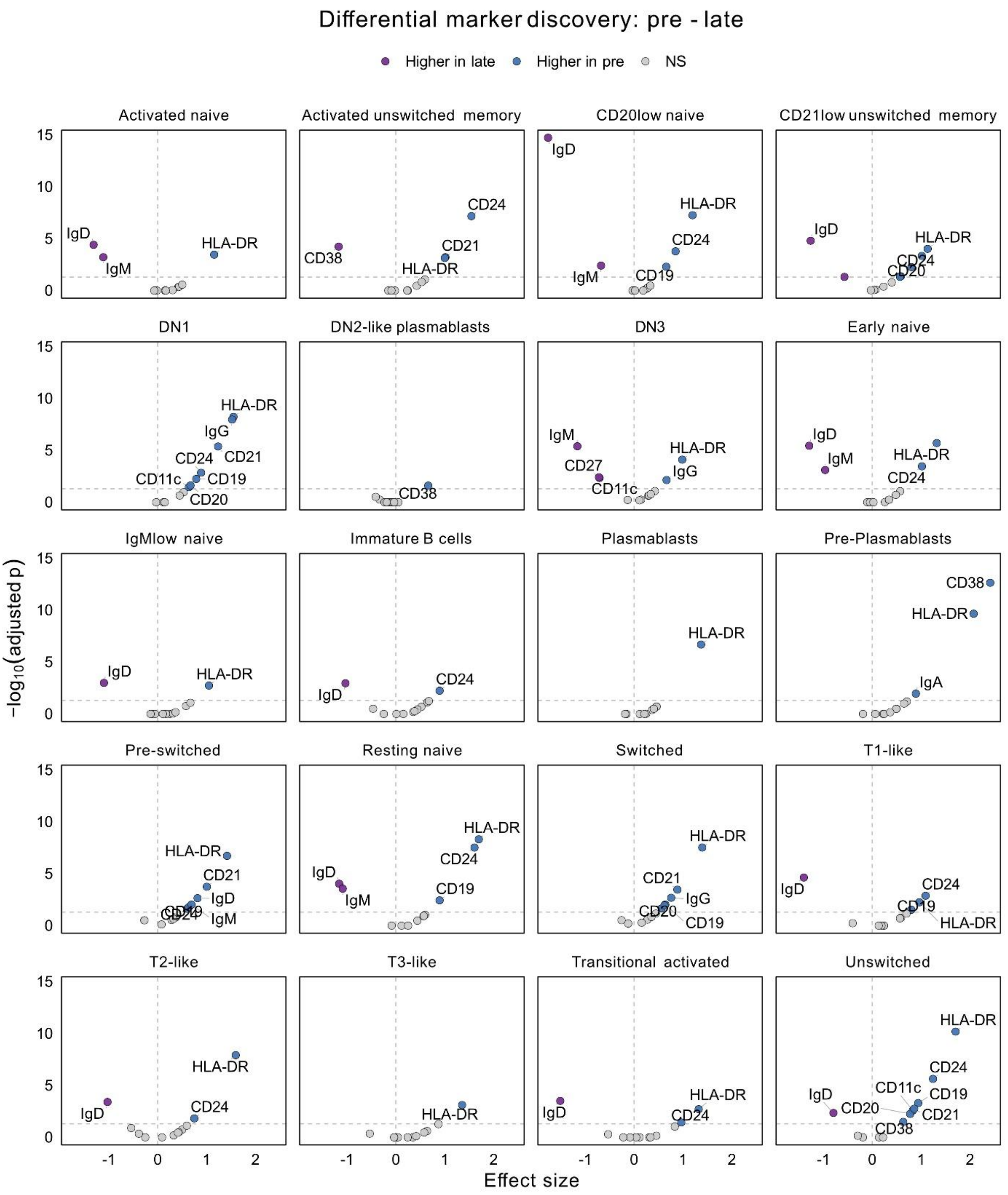


**Supplementary Fig. 4 | Differential marker discovery on peripheral B cells**

Faceted volcano plots showing differential flow cytometry marker staining intensity between pre-treatment and late time points across annotated B-cell subclusters. The x axis shows the estimated effect size for the pre versus late contrast, and the y axis shows -$\log_{10}$ Bonferroni-adjusted p value. Statistical testing was performed using linear mixed-effects models on sample-level mean marker staining intensity, with time point, marker and their interaction as fixed effects and patient/sample as a random intercept, followed by Bonferroni-adjusted pairwise contrasts.

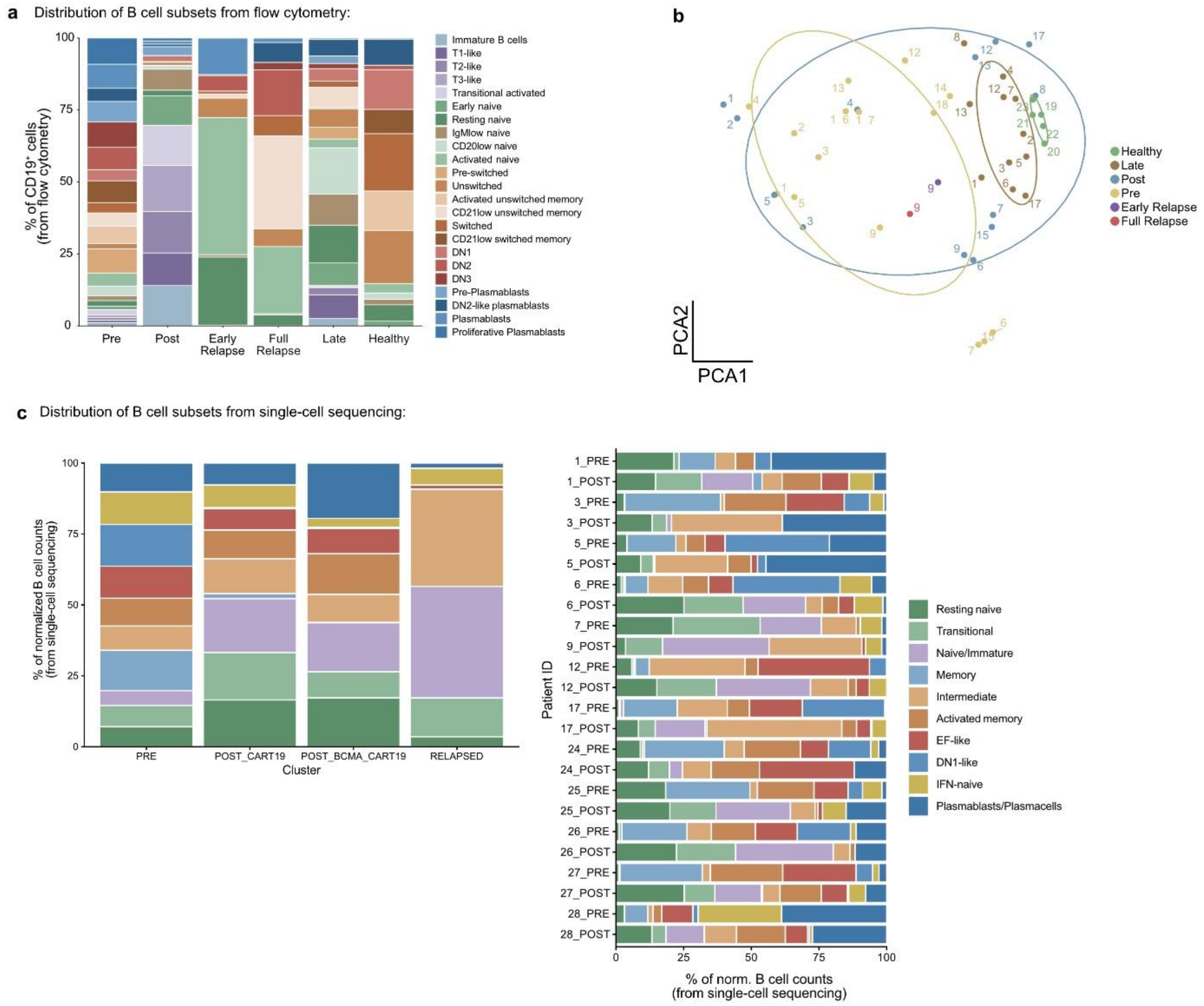


**Supplementary Fig. 5 | Flow cytometry and single-cell annotation of B cell subset composition**

**a**, Relative frequencies of flow cytometry-annotated B cell subsets across clinical time points. Stacked bars show the proportion of each B cell subset among CD19⁺ B cells in healthy controls and patients with SLE at baseline, post-treatment, late reconstitution and relapse-related time points. **b**, Principal component analysis of flow cytometry-derived B cell subset composition across samples. Points represent individual samples and are colored by clinical status. Early and late relapse samples are highlighted to show their relationship to baseline, post-treatment and healthy B cell states. **c**, Relative frequencies of scRNA-seq-annotated $CD19^{+}$ B cell states across clinical stages, including baseline, after CD19-CAR-T cell therapy, after CD19-BCMA dual CAR-T cell therapy and in relapse. Patient-level scRNA-seq B cell composition is additionally shown. Colors indicate annotated B cell states, and bars show the percentage of normalized $CD19^{+}$ cell counts assigned to each state. Norm., normalized.

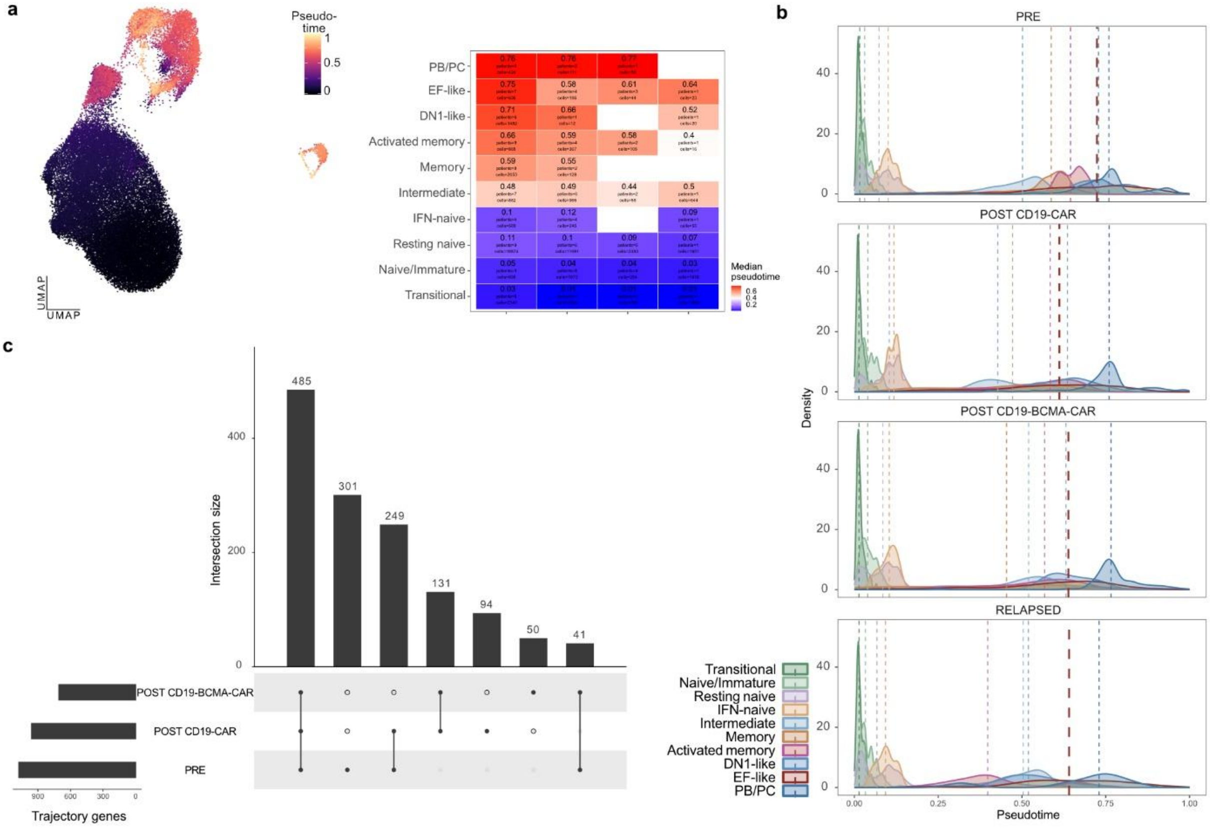


**Supplementary Fig. 6 | Pseudotime analysis of B cell maturation after CAR-T cell therapy**

**a**, Pseudotime ordering of scRNA-seq B cells across annotated B cell states. Left, pseudotime projected on UMAP. Right, heatmap showing median pseudotime values for each annotated B cell state across clinical stages, including baseline, after CD19 CAR-T cell therapy, after CD19/BCMA dual CAR-T cell therapy and relapse. **b**, Density plots that show the relative positioning of annotated B cell states along the inferred maturation trajectory at baseline, after CD19-CAR-T cell therapy, after CD19/BCMA dual CAR-T cell therapy and at relapse. Dashed vertical lines indicate median pseudotime values for each B cell state. The median value of EF-like B cells is highlighted in bold red. **c**, UpSet plot showing overlap of trajectory-associated genes identified at baseline, after CD19-CAR-T cell therapy and after CD19/BCMA dual CAR-T cell therapy as well as the absolute number of identified genes.

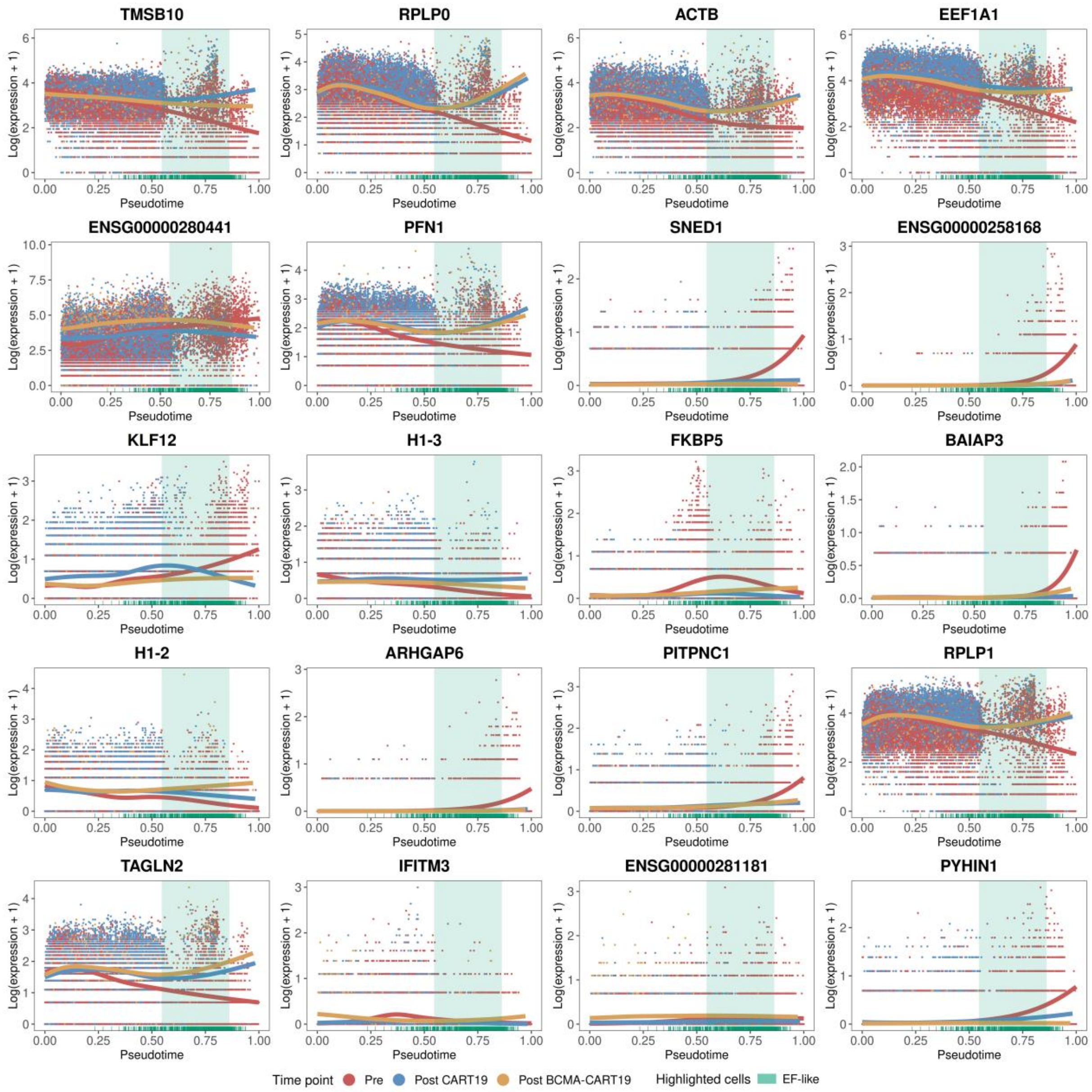


**Supplementary Fig. 7 | Top genes associated with the baseline B cell pseudotime trajectory**

Fitted expression dynamics of the top 20 PRE-specific pseudotime-associated genes along the inferred B-cell trajectory. Points show single-cell log-normalized expression, and smooth curves show fitted tradeSeq expression trends for PRE, POST_CART19 and POST_BCMA_CART19 conditions. The shaded region highlights the 10th–90th percentile pseudotime range of EF-like B cells.

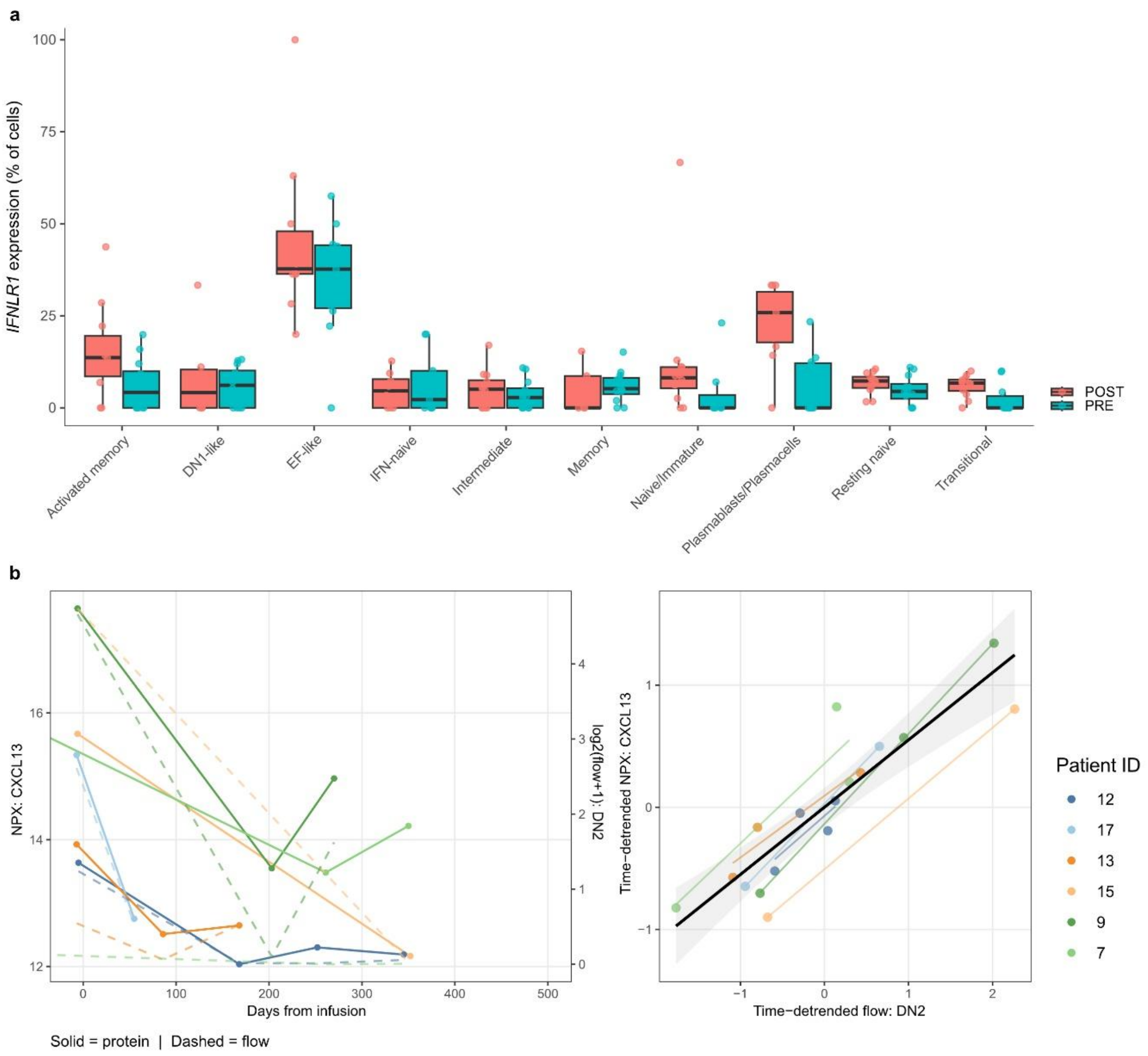


**Supplementary Fig. 8 | *IFNLR1* expression and coupling of serum CXCL13 with DN2 B cells**

**a**, scRNA-seq-based quantification of *IFNLR1* transcript detection across B cell states before therapy (PRE) and after B cell reconstitution (POST). Values indicate the percentage of cells within each cell type with detectable *IFNLR1* expression. Boxplots show the distribution across samples, with individual observations shown as dots, lines indicating the median and boxes showing the IQR. **b**, Representative time-adjusted longitudinal association between serum CXCL13 and flow cytometric DN2 B cell frequencies after CD19 CAR-T cell therapy. Left, matched longitudinal trajectories of serum CXCL13 protein abundance and DN2 frequencies are shown for individual patients (n = 6). Right, time-detrended values were correlated after adjustment for nonlinear time after infusion. The β coefficient, nominal p value and Benjamini-Hochberg-adjusted p value are shown. NPX, normalized protein expression. Flow, flow cytometry.

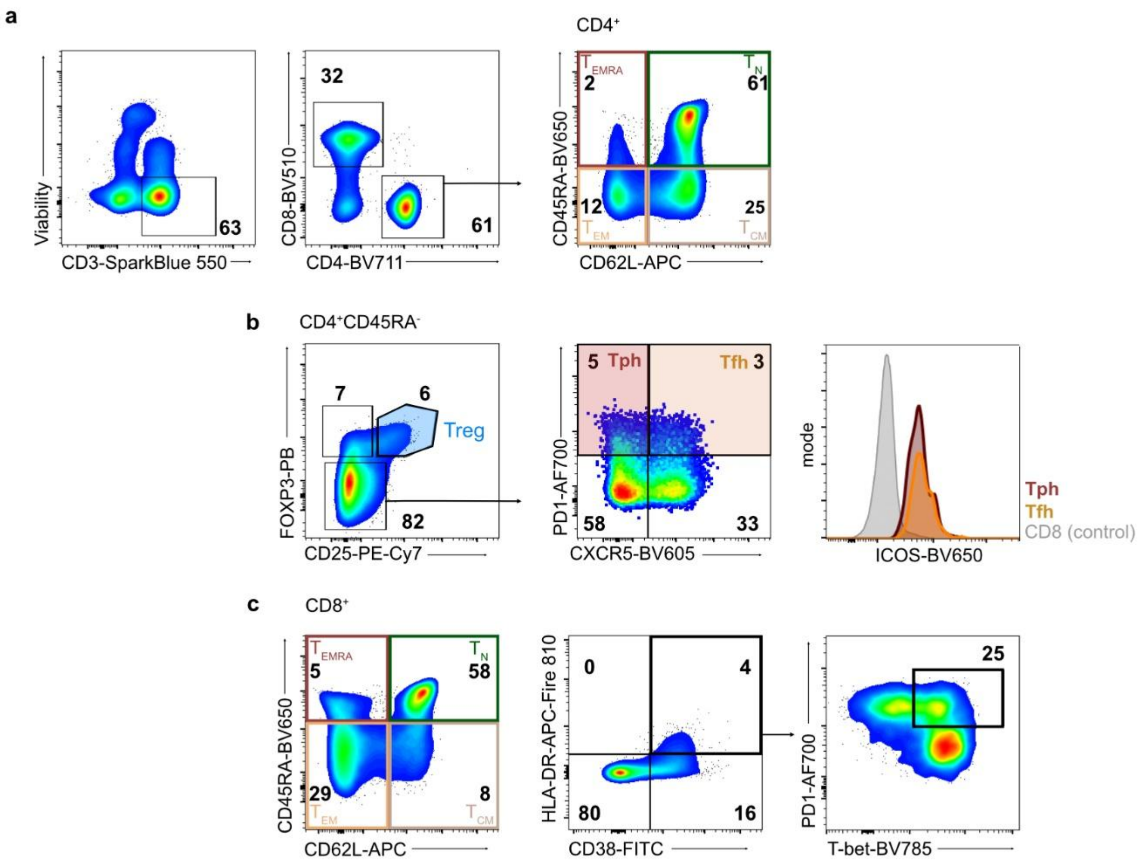


### Supplementary Fig. 9 │ Flow cytometry gating strategy for T cell subsets

Representative flow plots showing gating strategy to identify T cell subsets. Peripheral blood PBMC were stained with antibodies against live/dead fixable dye and doublets were excluded (not shown). **a**, Viable CD3$^+$ T cells were gated, followed by separation of CD4$^+$ and CD8$^+$ T cells. CD4$^+$ T cells were further classified using CD45RA and CD62L into naive T cells (TN), central memory T cells (TCM), effector memory T cells (TEM) and terminally differentiated effector memory CD45RA$^+$ T cells (TEMRA). **b**, Representative gating of CD4$^+$CD45RA$^-$ memory T cell subsets. Regulatory T cells were identified by CD25 and FoxP3 expression. T peripheral helper cells were defined as PD-1$^+$CXCR5$^-$ cells and T follicular helper cells as PD-1$^+$CXCR5$^+$ cells. ICOS expression is shown in Tph and Tfh cells, with CD8$^+$ T cells shown as control. **c**, Representative gating of CD8$^+$ T cell memory and activation states. CD8$^+$ T cells were classified using CD45RA and CD62L into naive, central memory, effector memory and terminally differentiated effector memory CD45RA$^+$ subsets as shown in a. Activated CD8$^+$ T cells were identified by CD38 and HLA-DR co-expression, followed by assessment of PD-1 and T-bet expression. Numbers indicate percentages of the respective parent population.

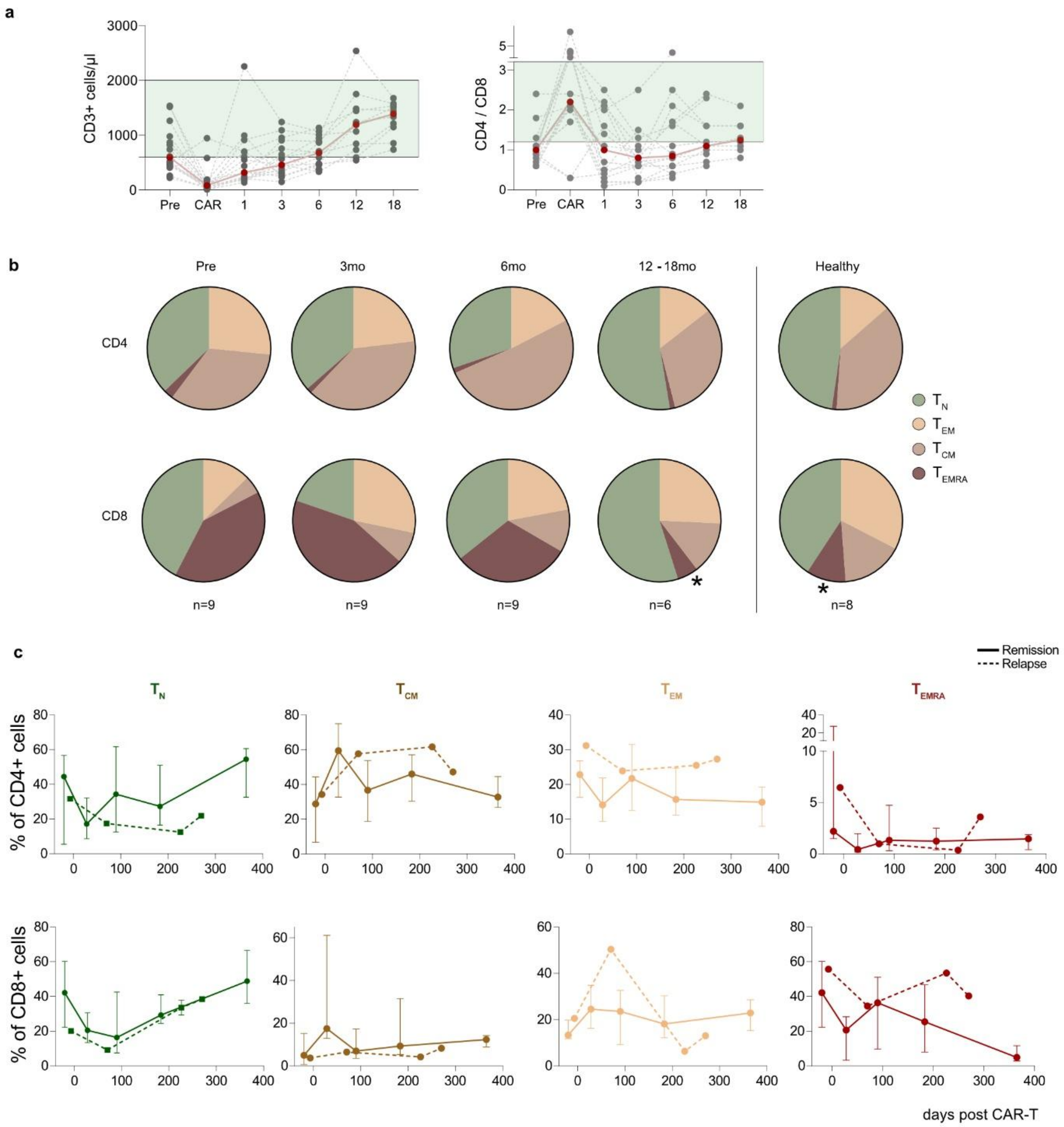


**Supplementary Fig. 10 │ Longitudinal remodeling of circulating T cell subsets after CAR-T cell therapy**

**a**, Longitudinal $CD3^+$ T cell counts and CD4/CD8 ratios after CAR-T cell therapy. Gray points and lines indicate individual patients; red points indicate the cohort median at each time point (n = 6-9 per timepoint). Green shading indicates the healthy reference range. **b**, Relative composition of $CD4^+$ and $CD8^+$ T cell subsets at baseline and during follow-up after CAR-T cell therapy, compared with healthy controls. Pie charts show parts-of-a-whole distributions calculated from median subset frequencies. Within $CD4^+$ and $CD8^+$ T cells, the subsets of naive T cells (TN), central memory T cells (TCM), effector memory T cells (TEM) and terminally differentiated effector memory T cells (TEMRA) were identified as shown in **Supplementary Fig. 9**. Sample numbers are indicated below the plots. Statistics: Kruskal-Wallis test and Dunn's correction within T cell subsets across timepoints. **c**, Longitudinal frequencies of $CD4^+$ and $CD8^+$ T cell memory subsets after CAR-T cell therapy. Solid lines show median values in patients maintaining remission, with error bars indicating the interquartile range. Dashed lines show the relapsed patient. Sample numbers correspond to those shown in **b**.

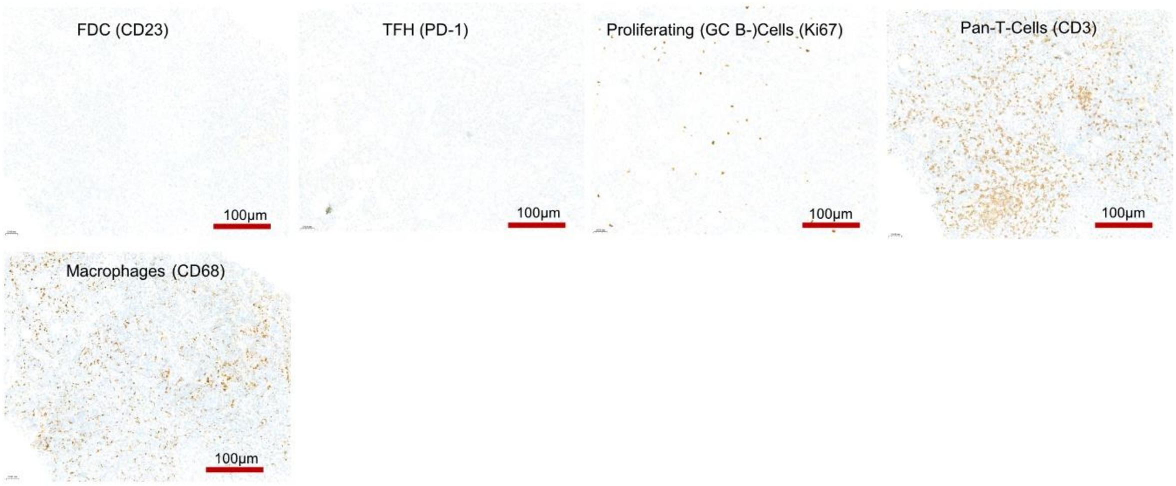


**Supplementary Fig. 11 | Additional lymph node immunohistochemistry during post-CAR-T B cell aplasia**

Representative immunohistochemical staining of inguinal lymph node tissue obtained from the relapsing patient at day 70 after CAR-T cell therapy during peripheral B cell aplasia. Sections were stained for follicular dendritic cells (CD23), PD-1$^{+}$ T follicular helper-like cells, proliferating cells (Ki67), pan-T cells (CD3) and macrophages (CD68). Scale bars, 100 µm. Where quantified, positive cells were assessed as cells per mm$^2$ using the same tissue-based quantification approach as for CD19, CD20, CD138, CD3 and CD68 staining.

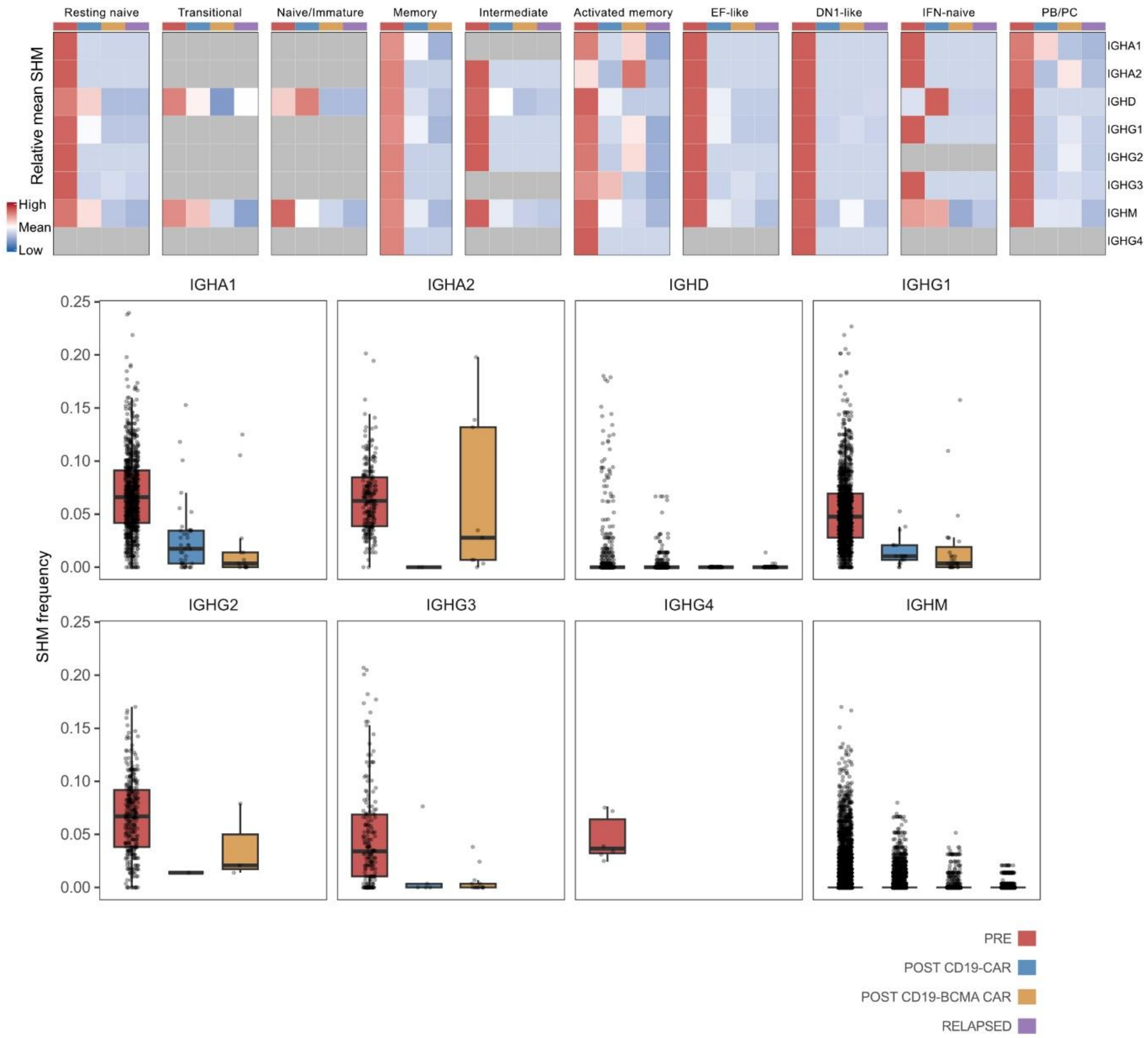


**Supplementary Fig. 12 | Somatic hypermutation across immunoglobulin isotypes after CAR-T cell therapy**

Heatmap showing relative mean somatic hypermutation frequency across immunoglobulin heavy-chain isotypes and annotated B cell states at baseline (PRE), after CD19 CAR-T cell therapy (POST CD19-CAR), after CD19/BCMA dual CAR-T cell therapy (POST CD19-BCMA-CAR) and at relapse (RELAPSED). Columns indicate clinical stages within each B cell state, and rows indicate heavy-chain isotypes. Color indicates relative mean SHM frequency; gray indicates insufficient or absent observations. Additionally, somatic hypermutation frequency stratified by immunoglobulin heavy-chain isotype across clinical stages is shown. Box plots show SHM frequency at baseline, after CD19 CAR-T cell therapy, after CD19/BCMA dual CAR-T cell therapy and at relapse.

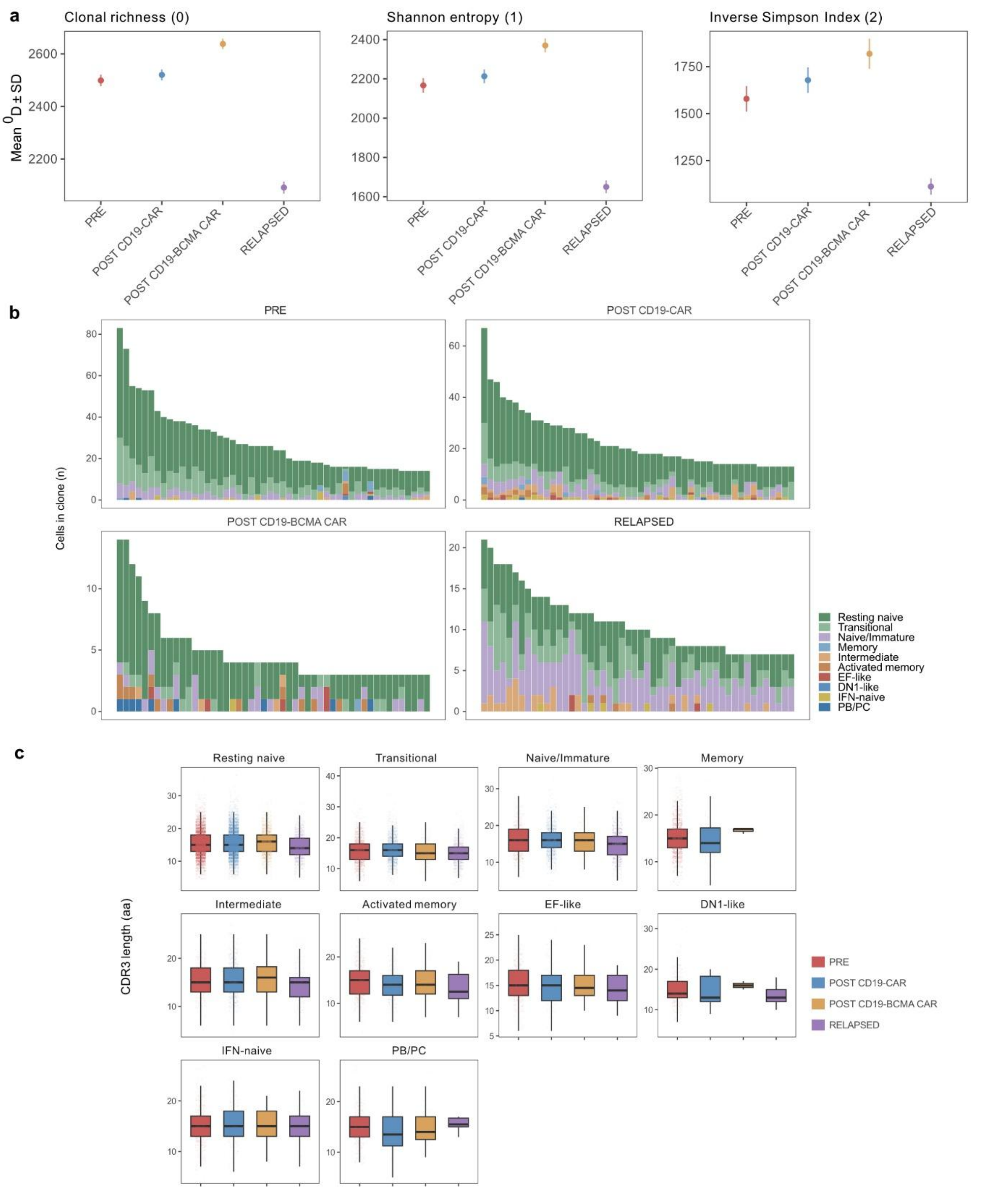


### Supplementary Fig. 13 | BCR clonal diversity and CDR3 length after CAR-T cell therapy

**a**, BCR repertoire diversity across clinical stages at baseline (PRE), after CD19 CAR-T cell therapy (POST CD19-CAR), after CD19-BCMA dual CAR-T cell therapy (POST CD19-BCMA-CAR) and at relapse (RELAPSED). Diversity is shown as clonal richness ($q = 0$), Shannon entropy ($q = 1$) and inverse Simpson index ($q = 2$). Points indicate mean values and error bars indicate SD. **b**, Composition of the largest BCR clones across clinical stages. Stacked bars show the top 50 clones ranked by clone size within each stage. Colors indicate the annotated B cell state contributing to each clone. **c**, CDR3 amino acid length across annotated B cell states and clinical stages. Box plots show the distribution of CDR3 length within each B cell subset at baseline, after CD19 CAR-T cell therapy, after CD19/BCMA dual CAR-T cell therapy and at relapse.

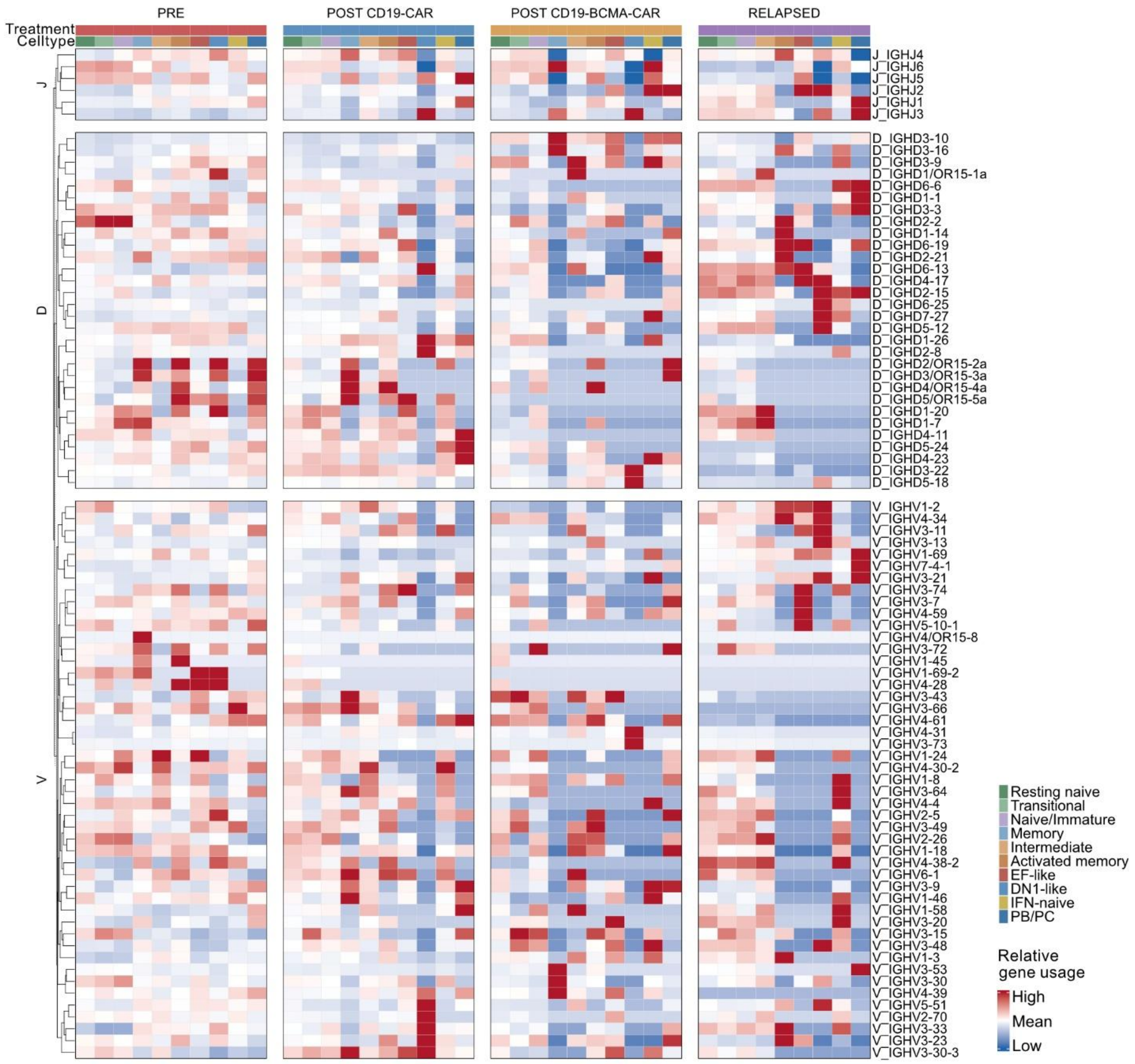


**Supplementary Fig. 14 | Immunoglobulin heavy-chain V(D)J gene usage across B cell states after CAR-T cell therapy**

Heatmap showing relative immunoglobulin heavy-chain variable (IGHV), diversity (IGHD) and joining (IGHJ) gene usage across annotated B cell states at baseline (PRE), after CD19 CAR-T cell therapy (POST CD19-CAR), after CD19/BCMA dual CAR-T cell therapy (POST CD19-BCMA-CAR) and at relapse (RELAPSED). Columns represent annotated B cell states within each clinical stage, and rows indicate individual IGHV, IGHD or IGHJ genes. Color indicates relative usage of each gene across cell states and clinical stages, with red indicating higher relative usage and blue indicating lower relative usage. Rows are hierarchically clustered within the IGHV, IGHD and IGHJ gene families.

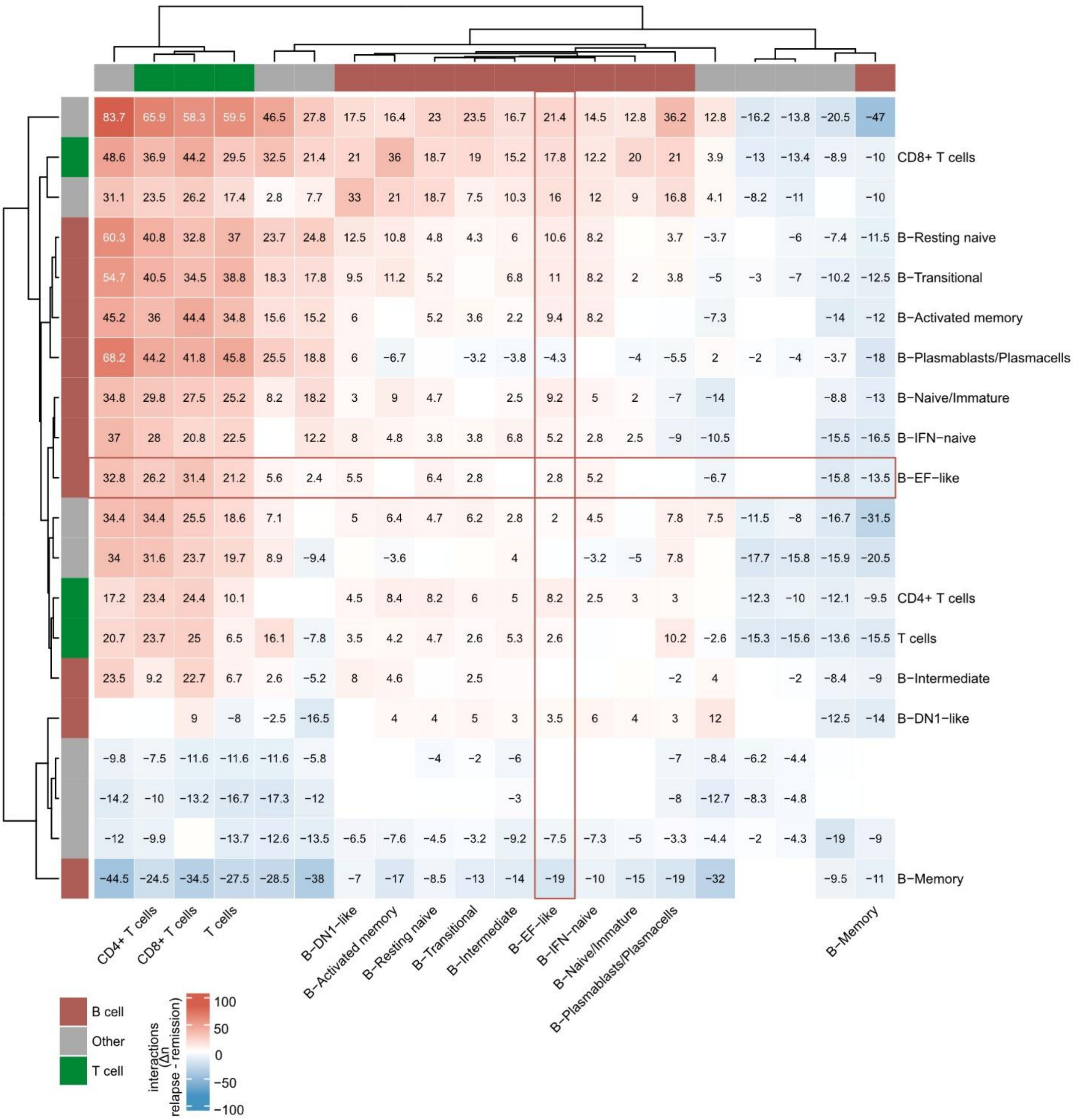


**Supplementary Fig. 15 | Differential cell-cell interaction density in relapse vs. remission**

Heat map showing the difference in the number (n) of significant ligand-receptor interactions between cell populations in relapse and post-CAR-T CD19 remission. Values and gradient represent Δ interaction numbers (relapse (n = 1) - mean CD19 remission (n = 10)) for interactions with an interaction score ≥ 0.9. Red indicates more interactions in relapse, whereas blue indicates more interactions in remission. Rows and columns represent the indicated PBMC cell populations, and top and side annotations indicate lineage class (B cell, T cell or other PBMC including mainly myeloid cells). Cell types were hierarchically clustered based on similarity of interaction changes across conditions. Interactions involving EF-like B cells are highlighted in a red frame.
